# Inferring transmission trees to guide targeting of interventions against visceral leishmaniasis and post-kala-azar dermal leishmaniasis

**DOI:** 10.1101/2020.02.24.20023325

**Authors:** Lloyd A. C. Chapman, Simon E. F. Spencer, Timothy M. Pollington, Chris P. Jewell, Dinesh Mondal, Jorge Alvar, T. Déirdre Hollingsworth, Mary M. Cameron, Caryn Bern, Graham F. Medley

**Affiliations:** Department of Global Health and Development and Centre for Mathematical Modelling of Infectious Disease, London School of Hygiene and Tropical Medicine, London WC1E 7HT, UK; Zeeman Institute, University of Warwick, Coventry CV4 7AL, UK; Centre for Health Informatics, Computing And Statistics, Lancaster University, Lancaster LA1 4YW, UK; International Centre for Diarrhoeal Disease Research Bangladesh, Dhaka 1212, Bangladesh; Drugs for Neglected Diseases initiative, Geneva 1202, Switzerland; Big Data Institute, Li Ka Shing Centre for Health Information and Discovery, University of Oxford, Oxford OX3 7LF, UK; Department of Disease Control, London School of Hygiene and Tropical Medicine, London WC1E 7HT, UK; Department of Epidemiology and Biostatistics, University of California San Francisco, San Francisco, CA 94158-2549, USA

**Author notes:** Author contributions: L.A.C.C., S.E.F.S., C.B. and G.F.M. designed research; L.A.C.C. and S.E.F.S. performed research; L.A.C.C., S.E.F.S., T.M.P., C.P.J., D.M., J.A., T.D.H., M.M.C., C.B. and G.F.M. contributed new reagents/analytic tools; L.A.C.C. and S.E.F.S. analysed data; L.A.C.C., S.E.F.S., C.B. and G.F.M. wrote the paper. The authors declare no conflict of interest.

**Keywords:** visceral leishmaniasis, post-kala-azar dermal leishmaniasis, spatiotem-transmission, transmission tree, Bayesian inference

## Abstract

Understanding of spatiotemporal transmission of infectious diseases has improved significantly in recent years. Advances in Bayesian inference methods for individual-level geo-located epidemiological data have enabled reconstruction of transmission trees and quantification of disease spread in space and time, while accounting for uncertainty in missing data. However, these methods have rarely been applied to endemic diseases or ones in which asymptomatic infection plays a role, for which novel estimation methods are required. Here, we develop such methods to analyse longitudinal incidence data on visceral leishmaniasis (VL), and its sequela, post-kala-azar dermal leishmaniasis (PKDL), in a highly endemic community in Bangladesh. Incorporating recent data on infectiousness of VL and PKDL, we show that while VL cases drive transmission when incidence is high, the contribution of PKDL increases significantly as VL incidence declines (reaching 55% in this setting). Transmission is highly focal: *>*85% of mean distances from inferred infectors to their secondary VL cases were *<*300m, and estimated average times from infector onset to secondary case infection were *<*4 months for 90% of VL infectors, but up to 2.75yrs for PKDL infectors. Estimated numbers of secondary VL cases per VL and PKDL case varied from 0-6 and were strongly correlated with the infector’s duration of symptoms. Counterfactual simulations suggest that prevention of PKDL could have reduced VL incidence by up to a quarter. These results highlight the need for prompt detection and treatment of PKDL to achieve VL elimination in the Indian subcontinent and provide quantitative estimates to guide spatiotemporally-targeted interventions against VL.

**Significance Statement:** Although methods for analysing individual-level geo-located disease data have existed for some time, they have rarely been used to analyse endemic human diseases. Here we apply such methods to nearly a decade’s worth of uniquely detailed epidemiological data on incidence of the deadly vector-borne disease visceral leishmaniasis (VL) and its secondary condition, post-kala-azar dermal leishmaniasis (PKDL), to quantify the spread of infection around cases in space and time by inferring who infected whom, and estimate the relative contribution of different infection states to transmission. Our findings highlight the key role long diagnosis delays and PKDL play in maintaining VL transmission. This detailed characterisation of the spatiotemporal transmission of VL will help inform targeting of interventions around VL and PKDL cases.

---

Spatiotemporal heterogeneity in incidence is a hallmark of infectious diseases. Insight into this heterogeneity has increased considerably in recent years due to greater availability of geo-located individual-level epidemiological data and the development of sophisticated statistical inference methods for partially observed transmission processes (1–6). These methods have been developed for epidemics, in which the immune status of the population is known, and for diseases with a short time course that are relatively easily diagnosed, such as measles, influenza, and foot-and-mouth disease (3, 4, 7). Here, we extend these methods to a slowly progressing endemic disease of humans in which asymptomatic infection plays an important role.

We analyse detailed longitudinal individual-level data on incidence of visceral leishmaniasis (VL), and its sequela, postkala-azar dermal leishmaniasis (PKDL), in a highly endemic community in Fulbaria, Bangladesh (8). VL, also known as kala-azar, is a lethal sandfly-borne parasitic disease targeted for elimination as a public health problem (*<*1 case/10,000 people/year at subdistrict/district level depending on the country) in the Indian subcontinent (ISC) by 2020 (9). PKDL is a non-lethal skin condition that occurs after treatment for VL in 5–20% of cases in the ISC, and less frequently in individuals who report no history of prior VL (8, 10). It is characterised by skin lesions of differing severity and parasite load, ranging from macules and papules (least severe, lowest load) to nodules (most severe, highest load) (11). We estimate the relative contributions of different disease states (VL, PKDL and asymptomatic infection) to transmission, and quantify the rate of spread of infection around infected individuals in space and time by reconstructing transmission trees. Our analysis provides insight into the spatiotemporal spread of visceral leishmaniasis as well as quantitative estimates that can guide the targeting of interventions, such as active case detection and indoor residual spraying (IRS) of insecticide, around VL

PKDL cases are believed to play a role in transmission of VL as historical and recent xenodiagnosis studies have shown that all PKDL forms are infectious towards sandflies (11–13), and a 1992 study in West Bengal, India, suggested that PKDL cases are capable of initiating a VL outbreak in a susceptible community (14). Furthermore, PKDL cases typically have long durations of symptoms before treatment and often go undiagnosed as the disease is not systemic (15–17). While VL incidence has declined considerably throughout the ISC since 2011 (by *>*85%, from *∼* 37,000 cases in 2011 to *∼* 4,700 in 2018) (18, 19), reported numbers of PKDL diagnoses increased from 590 in 2012 to 2,090 in 2017 before falling to 1,363 in 2018 (19, 20). PKDL has therefore been recognised as a major potential threat to the VL elimination programme in the ISC (10), which has led to increased active PKDL case detection. Nevertheless, the contribution of PKDL to transmission in field settings still urgently needs to be quantified.

Although the incidence of asymptomatic infection is 4 to 17 times higher than that of symptomatic infection in the ISC (21), the extent to which asymptomatic individuals contribute to transmission is still unknown (22, 23). What is clear is that asymptomatic infection plays a role in transmission through generating herd immunity, since a significant proportion of asymptomatically infected individuals develop protective cell-mediated immunity against VL following infection, as measured by positivity on the leishmanin skin test (LST) (24–27). Several studies have shown that asymptomatic infection is spatiotemporally clustered (25, 28), and therefore immunity is also likely to be spatially clustered, but so far no transmission models have accounted for this (23). Since most surveillance data and data from epidemiological studies does not contain information about numbers of asymptomatically infected individuals over space and time (e.g. from longitudinal serological testing), accounting for the role of asymptomatic infection in transmission at the individual level represents a substantial missing data problem. The endemic nature of the disease and high asymptomatic infection potential mean that it is necessary to infer initial infection statuses for individuals without symptomatic disease, unlike for many epidemic diseases where individuals can be assumed to be susceptible or are known to have been vaccinated. Coupled with the long and variable incubation period of VL (lasting anywhere between weeks and years but typically 2-6 months (29, 30)) and lack of data on the flight range of the *P. argentipes* sandfly vector, these factors make inference of spatiotemporal transmission of VL particularly challenging.

By combining data from a recent xenodiagnosis study in Bangladesh (11) with the geo-located data on incidence and duration of symptoms of VL and different forms of PKDL from the community study in Bangladesh, and fitting it to an individual-level spatiotemporal VL transmission model, this study provides the first detailed insight into the changing roles of VL, PKDL, asymptomatic infection and immunity in transmission over the course of an epidemic, and the first estimates of numbers of secondary cases and infections generated by individual VL and PKDL cases. The Bayesian data augmentation framework that we develop in order to fit the model accounts for the unobserved infection times of VL cases, the missing data on asymptomatic infections, individuals’ unobserved initial infection statuses, migration of individuals and uncertainty in infection sources, and could be readily adapted to analyse spatiotemporal transmission of other endemic diseases in which asymptomatic infection plays a hidden role.

## Study Data

We analyse detailed demographic and disease data on 24,781 individuals living in 5,118 households in 19 paras (hamlets) situated in two large clusters in a 12km *×* 12km area in Fulbaria upazila, Mymensingh district, Bangladesh from 2002-2010 (Fig. 1A). The data from this study are fully described elsewhere (8, 31). Briefly, month of onset of symptoms, treatment, relapse, and relapse treatment were recorded for VL cases and PKDL cases with onset between 2002 and 2010 (retrospectively for cases with onset before 2007), and year of onset was recorded for VL cases with onset before 2002. There were 1018 VL cases and 190 PKDL cases with onset between January 2002 and December 2010 in the study area, and 413 VL cases with onset prior to January 2002.

Over the whole study area, VL incidence followed an epidemic wave, increasing from approximately 40 cases/10,000/yr in 2002 to *∼*90 cases/10,000/yr in 2005 before declining to *<*5 cases/10,000/yr in 2010 (Fig. 1B). PKDL incidence followed a similar pattern but lagging VL incidence by roughly 2yrs, peaking at 30 cases/10,000/yr in 2007. However, VL and PKDL incidence varied considerably across paras (average para-level incidences: VL 18–124 cases/10,000/yr, PKDL 0-31 cases/10,000/yr, Table S5) and time (range of annual para-level incidences: VL 0–414 cases/10,000/yr, PKDL 0–120 cases/10,000/yr, Fig. S15).

## Results

### Model Comparison

Different versions of the spatiotemporal transmission model described in *Materials and Methods*, in which decrease in infection risk with distance from an infectious individual is characterised by an exponentially decaying spatial kernel function, were fitted to the data. These comprised models with and without extra within-household transmission (over and above that from being at zero distance from an infectious individual) and with different pre-symptomatic and asymptomatic relative infectiousness. Models with additional within-household transmission fitted the data significantly better than those without according to the Deviance Information Criterion (DIC) (32), and for these models fit improved with increasing pre-symptomatic and asymptomatic infectiousness, such that the best-fitting model had additional within-household transmission and assumed 2% relative infectiousness of pre-symptomatic and asymptomatic individuals. Here we present the results from this model (see *SI Text* for results from the other models).

**Fig. 1.**
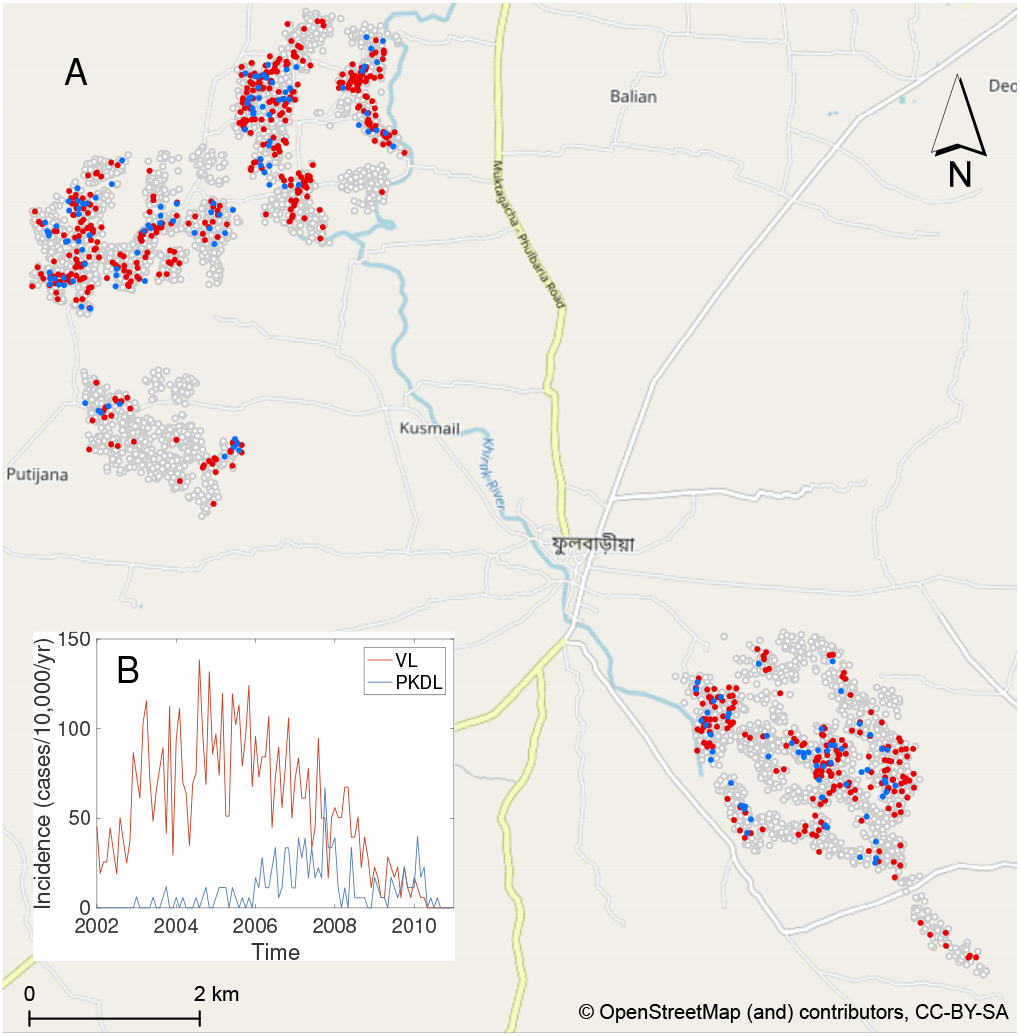
(A) Map of the study area showing the households that had VL cases (red), PKDL cases (blue), and no cases (white with grey outline) with onset between 2002 and 2010. Household locations jittered slightly to protect patient anonymity. (B) Observed incidence of VL and PKDL for the whole study area by month of onset, 2002-2010.

### Parameter Estimates

We estimated the transmission model parameters and unobserved data using the Markov Chain Monte Carlo (MCMC) algorithm described in *Materials and Methods* and *SI Text*. The posterior distributions obtained for the model parameters are shown in Fig. S6 and the corresponding posterior modes and 95% credible intervals (CI) are given in Table 1.

**Table 1.** Transmission parameter estimates from the spatiotemporal model

| Parameter | Mode | 95% CI* |
| --- | --- | --- |
| Risk of developing VL<sup>†</sup> (month<sup>-1</sup>) if living in the same household as a: |  |  |
| - VL case | 0.018 | (0.014, 0.024) |
| - PKDL case: |  |  |
| - macular/papular | 0.009 | (0.007, 0.013) |
| - plaque<sup>‡</sup> | 0.016 | (0.012, 0.022) |
| - nodular | 0.023 | (0.018, 0.031) |
| - asymptomatic individual | 0.00037 | (0.00027, 0.00049) |
| Risk of asymptomatic infection<sup>†</sup> (month<sup>-1</sup>) if living in the same household as a: |  |  |
| - VL case | 0.101 | (0.075, 0.132) |
| - PKDL case: |  |  |
| - macular/papular | 0.052 | (0.040, 0.070) |
| - plaque<sup>‡</sup> | 0.092 | (0.068, 0.118) |
| - nodular | 0.126 | (0.096, 0.166) |
| - asymptomatic individual | 0.0021 | (0.0016, 0.0028) |
| Risk of developing VL from background transmission (month<sup>-1</sup>) | 6.9 × 10<sup>-5</sup> | (3.8 × 10<sup>-5</sup>, 11 × 10<sup>-5</sup>) |
| Risk of asymptomatic infection from background transmission (month<sup>-1</sup>) | 3.9 × 10<sup>-4</sup> | (2.2 × 10<sup>-4</sup>, 6.2 × 10<sup>-4</sup>) |
| Decrease in risk of infection with distance from an infectious individual (per 100m)<sup>§</sup> | 58.0% | (52.7%, 63.8%) |
| Hazard ratio for increase in infection risk from living in the same household as an infectious individual compared to living just outside | 12.0 | (8.3, 16.7) |

\* CI = credible interval, calculated as the 95% highest posterior density interval
<sup>†</sup> risk of subsequent VL/asymptomatic infection if susceptible
<sup>‡</sup> based on assumed infectiousness
<sup>§</sup> in the absence of background transmission and relative to living directly outside the case household.

Based on the relative infectiousness of VL and the different types of PKDL from the xenodiagnostic data, in the absence of any other sources of transmission, the estimated probability of being infected and developing VL if living in the same household as a single symptomatic individual for 1 month following their onset was 0.018 (95% CI: 0.013, 0.024) for VL and ranged from 0.009 to 0.023 (95% CIs: (0.007,0.013)–(0.018, 0.031)) for macular/papular PKDL to nodular PKDL. Living in the same household as a single asymptomatic individual, the monthly risk of VL was only 0.00037 (95% CI: 0.00027, 0.00049), if asymptomatic individuals are 2% as infectious as VL cases.

The risk of infection if living in the same household as an infectious individual was estimated to be more than 10 times higher than that if living directly outside the household of an infectious individual (hazard ratio = 12.0), with a 95% CI well above 1 (8.3, 16.7). The estimated spatial kernel (Fig. S16) around each infectious individual shows a relatively rapid decay in risk with distance *outside* their household, the risk of infection halving over a distance of 84m (95% CI: 71, 99m).

### Contribution of PKDL and Asymptomatic Infection to Transmission

We assess the contribution of different infectious groups to transmission in terms of their relative contribution to the transmission experienced by susceptible individuals (Fig. 2A and Fig. S17). The contribution of VL cases was fairly stable at around 75% from 2002 to the end of 2004 before decreasing steadily to 0 at the end of the epidemic, while the contribution of PKDL cases increased from 0 in 2002 to *∼* 73% in 2010 (95% CI: 63, 80%) (Fig. S17). Only a small proportion of the total infection pressure on susceptible individuals, varying between 9% and 14% over the course of the epidemic, was estimated to have come from asymptomatic and pre-symptomatic individuals.

**Fig. 2.**
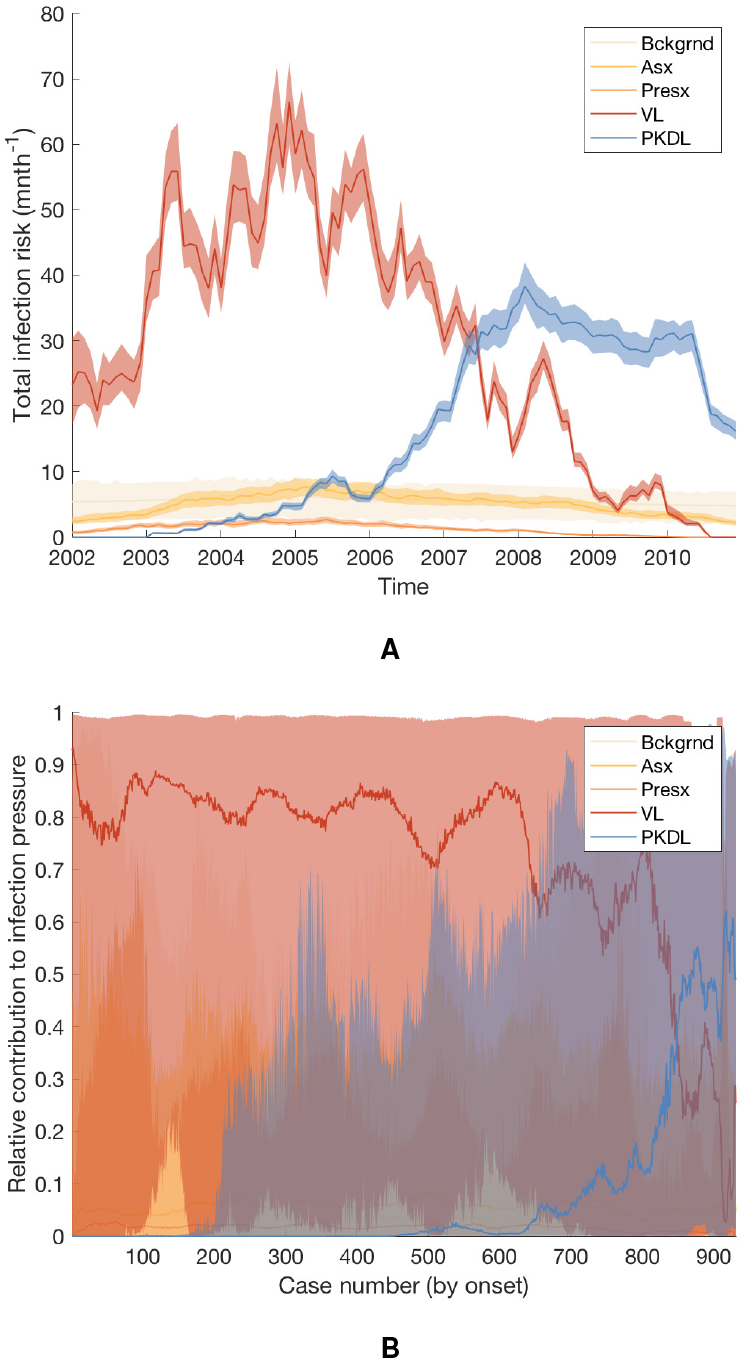
Contributions of background transmission, asymptomatic individuals, pre-symptomatic individuals, VL cases and PKDL cases to (A) the total risk of new infections, and (B) the individual infection pressures on VL cases at their infection times (in relative terms). Note that time is non-linear in (B) since cases are ordered by their onset time. Solid lines show modes in (A), medians in (B); shaded regions show 95% CIs. The relative contribution of PKDL to the infection pressures on the 7 VL cases with onset in 2010 in (B) is lower than to the infection pressure on susceptible individuals in 2010 in (A) since the 2010 VL cases all had onset before May and were therefore most likely infected in 2009 when the relative contribution of VL was higher.

Fig. 2B shows the breakdown of the individual infection pressures on VL cases at their infection times, and indicates that the contribution of PKDL to these infection pressures grew from 0% at the start of the epidemic to approximately 55% (95% CI: 2, 92%) for the cases with onset in 2010. Unsurprisingly, given the uncertainty in the infection times of the VL cases, the credible intervals for the relative contributions of each infection source to the infection pressures on the cases at their infection times are very broad.

### Reconstructing the Transmission Tree

By sampling 1,000 transmission trees from the joint posterior distribution of the transmission parameters and the unobserved data (as described in *Materials and Methods*), we can build a picture of the most likely source of infection for each case and how infection spread in space and time. Fig. 3 shows the transmission tree at different points in time in part of the south-east cluster of villages. Early in the epidemic and at its peak (Figures 3A and 3B), most new infections were due to VL cases. Towards the end of the epidemic, some infections were most likely due to PKDL cases and there was some saturation of infection around VL cases (Fig. 3C). The inferred patterns of transmission suggest that disease did not spread radially outward from index cases over time, but instead made a combination of short and long jumps around cases with long durations of symptoms and households with multiple cases.

**Fig. 3.**
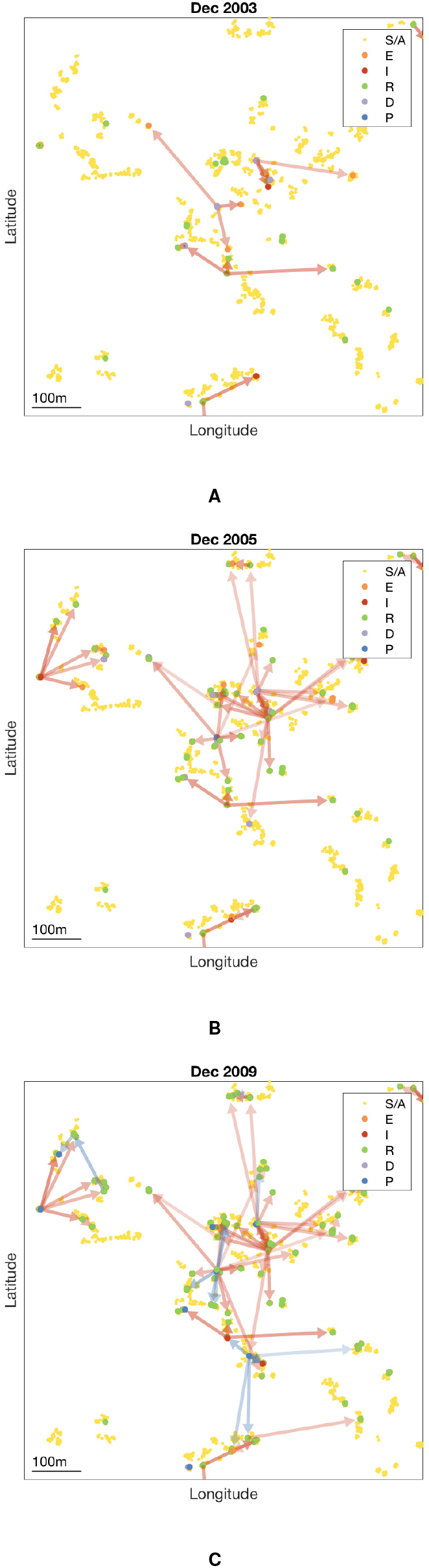
Inferred transmission tree in part of the south-east cluster of villages at different stages of the epidemic: (A) Dec 2003, (B) Dec 2005, and (C) Dec 2009. Dots show individuals coloured by their infection state (see key). Arrows show the most likely source of infection for each case infected up to that point in time over 1,000 sampled transmission trees, and are coloured by the type of infection source and shaded according to the proportion of trees in which that individual was the most likely infector (darker shading indicating a higher proportion). Asymptomatic infections are not shown for clarity. S/A = susceptible or asymptomatic, E = pre-symptomatic, I = VL, R = recovered, D = dormantly infected, P = PKDL (see *SI Text*). GPS locations of individuals are jittered slightly so that individuals from the same household are more visible. An animated version showing all months is provided in SI movie 1.

### Transmission Distances and Times

Having reconstructed a set of samples of the transmission tree as described above, we can use them to calculate the mean distance from each VL/PKDL infector to their VL-case infectees and the mean times between their onset and the infections of their VL-infectees, to assess how far and how quickly interventions need to be performed around VL and PKDL cases. Fig. 4A shows that the mean distances to VL-infectees for VL and PKDL cases are mostly within 500m but tend to be greater for PKDL cases (median 217m, inter-quartile range (IQR): 151, 310m) than VL cases (median 163m, IQR: 102, 233m), reflecting the fact that around PKDL cases there has typically already been considerable transmission from prior VL and therefore development of immunity in asymptomatically infected individuals. However, the mean times between infector onset and VL-infectee infections are much greater for PKDL cases (median 5.6 months, IQR: 3.0, 9.7 months) than VL cases (median 1.9 months, IQR: 1.4, 2.7 months) (Fig. 4B). Thus, whilst a similar intervention radius around new VL/PKDL cases of *∼* 500m may be sufficient to capture most secondary VL cases, the time window within which interventions need to be performed to prevent secondary cases is much narrower for VL cases than PKDL cases.

**Fig. 4.**
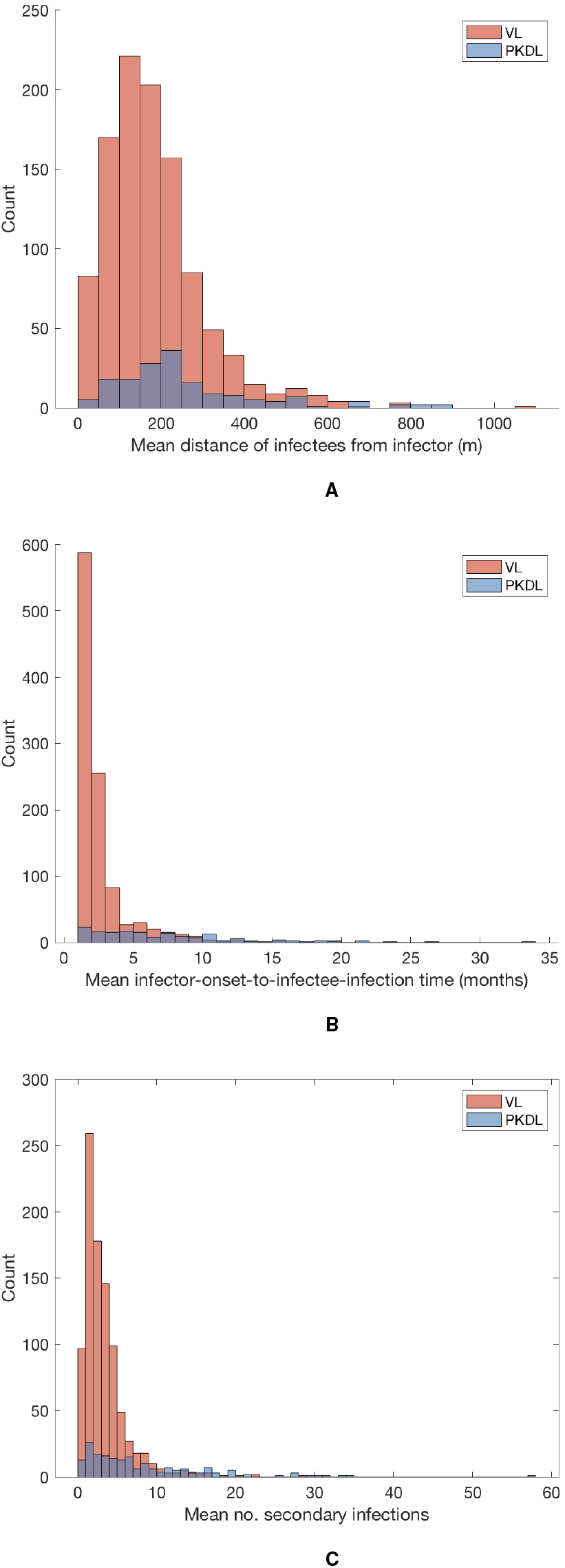
(A) Distributions of mean numbers of secondary infections per VL and PKDL case. (B) Mean distances from VL and PKDL infectors to their VL infectees. (C) Mean times from symptom onset of VL and PKDL infectors to the infections of their VL infectees.

### Numbers of Secondary Infections

Since we infer the unobserved infection times of VL cases and asymptomatic individuals as part of the MCMC algorithm, we can calculate the probability that each individual was infected by another individual conditional on their estimated infection month. Using these probabilities, we can then estimate the numbers of secondary infections generated by each infectious individual.

The mean numbers of secondary infections per VL case and per PKDL case (Fig. 4C) show large variation, ranging from 0.4 to 28.3 for VL and 0.2 to 57.3 for PKDL (see Fig. S18 for the posterior distributions of the number of secondary infections generated by each VL and PKDL case), and are overdispersed, with shape parameters for fitted gamma distributions of 1.98 (95% confidence interval: 1.82, 2.15) and 1.21 (95% confidence interval: 1.01, 1.44) respectively. This indicates that some cases generate far more secondary infections than others, a phenomenon known as ‘super-spreading’, which has been observed for a variety of diseases (33, 34), and hypothesised for VL (22, 35). The estimated mean numbers of secondary infections for asymptomatic individuals are much lower, ranging from 0 to 0.87. Whilst the numbers of secondary infections for VL and PKDL may seem high, we note that they are the number of new pre-symptomatic *and* asymptomatic infections generated by each case, and that only approximately 1 in 7 new infections were estimated to have led to VL (30), so the estimated numbers of secondary *VL* cases per case are much lower (Fig. S19A).

As expected, the mean numbers of secondary infections generated by infectious individuals are strongly positively correlated with their durations of infectiousness (Fig. S19B). In particular, many PKDL cases had very long durations of symptoms (*>*1yr) and generated large numbers of secondary infections (*>*5).

The median effective reproduction number—the average number of secondary infections generated by individuals who became infectious in a given month, which must be above 1 for the disease to persist—decreased over the course of the epidemic (Fig. S19C), from being mostly above 1 (range: 0.4, 3.7) in 2003-2006 to below 1 in 2007-2010.

### Impact of Preventing/Limiting PKDL

To investigate the potential impact of stopping PKDL from occurring or reducing the duration of infectiousness of PKDL cases on incidence of VL, we created a simulation version of the transmission model and used the parameter estimates and inferred initial statuses of individuals obtained from the MCMC algorithm to run counterfactual simulations of the epidemic in the study area (see *Materials and Methods* and *SI Text* for further details). Based on these simulations, if there had been no PKDL, the total number of VL cases from 2002–2010 would have been 25% lower (95% CI: 5, 42%) (see Fig. S20 and Table S6 for the para-level impact). This is the hypothetical maximum proportion of VL cases that could be averted by preventing any PKDL, e.g. if there was a vaccine available that could prevent progression to PKDL (36). However, even if the mean duration of infectiousness of PKDL had only been halved (from 17.8 months to 8.9 months)—which represents a more realistically achievable target in the near future through improved active case detection—the simulations suggest the total number of VL cases would have been 9% lower (95% CI: −15, 29%).

## Discussion

This study represents the first attempt to estimate the contribution of PKDL to transmission of VL accounting for spatiotemporal clustering of VL and PKDL and unobserved asymptomatic infection. It is also the first study to combine infectiousness data from xenodiagnostic studies with geo-located VL and PKDL incidence data, and to use this to reconstruct transmission trees of the spread of VL through a community and estimate individual-level numbers of secondary infections.

Our results support the conclusion that PKDL poses a significant threat to the VL elimination programme in the Indian subcontinent. Whilst VL cases drive transmission when VL incidence is high during the peak years of an epidemic, the contribution of PKDL to transmission increases as VL incidence decreases and PKDL prevalence increases in the downward phase of an epidemic. This mirrors the current situation in Bangladesh and India, where VL incidence has been decreasing since 2011 (18, 19), but reported numbers of PKDL cases suggest PKDL prevalence is higher than VL prevalence in some areas (19, 37).

In the study area in Bangladesh the contribution of PKDL (in terms of contribution to new symptomatic infections) grew from close to 0% in the upward phase of the epidemic in 2002-2005, to approximately 55% at the end of the epidemic in 2010. In light of the current low VL incidence and considerable numbers of PKDL cases being reported in much of the Indian subcontinent, this suggests that measures need to be taken to ensure all PKDL cases are detected and treated in order to maintain reduced transmission. This will require improvements in both active PKDL case detection, e.g. through comprehensive long-term follow-up of VL cases, and diagnostic tests and algorithms and treatment regimens for PKDL (17, 38, 39).

There is considerable heterogeneity in the estimated contribution of individual VL cases and PKDL cases to transmission in terms of the numbers of secondary infections they generate, which is chiefly driven by variation in their onset-to-recovery times (Fig. S19B). As expected, individuals with long onset-to-recovery times contribute most to new infections, acting as super-spreaders who generate many times more infections than the average case. These individuals play an important role in maintaining transmission of VL—keeping the effective reproduction number above 1—as the average number of secondary VL cases (the main drivers of transmission) generated by each VL/PKDL case is typically less than 1 (Fig. S19A). The times after onset of symptoms in the infector at which secondary VL cases become infected are typically longer for PKDL infectors than for VL infectors (Fig. 4B), due to their longer durations of infection and generally lower infectiousness, so there is greater opportunity to intervene to prevent onward transmission from PKDL cases. Model simulations suggest that incidence of VL could be reduced by faster detection and treatment of PKDL cases, e.g. by nearly 10% by halving the average duration of PKDL infectiousness, and by a quarter if it were possible to prevent PKDL altogether.

The spatiotemporal patterns of transmission inferred from reconstructing the transmission tree suggest that infection makes both short and long jumps in space within each infection generation rather than spreading radially outward from index cases in a wave. This is consistent with findings from a spatial analysis of occurrence of VL cases around index cases in Muzaffarpur, Bihar, India (40), which found a combination of short and long distances (from tens to hundreds of metres) from the closest index case for secondary VL cases diagnosed close together in time. Considering that index cases are often detected after a longer delay than subsequent cases and there will be some delay in mounting a reactive intervention, such as active case detection and/or targeted IRS around the index case(s), interventions will need to be applied in a large radius (up to 500m) around index cases to be confident of capturing all secondary cases and limiting transmission.

Our results demonstrate the importance of accounting for spatial clustering of infection and disease when modelling VL transmission. Previous VL transmission dynamic models (23, 41–43) have significantly overestimated the relative contribution of asymptomatic infection to transmission (as up to 80%), despite assuming asymptomatic individuals are only 1-3% as infectious as VL cases, by treating the population as homogeneously mixing, such that all asymptomatic individuals can infect all susceptible individuals via sandflies. In reality, asymptomatic individuals do not mix homogeneously with susceptible individuals as they are generally clustered together around or near to VL cases (25, 28), who are much more infectious and therefore more likely to infect susceptible individuals around them, even if they are outnumbered by asymptomatic individuals. Asymptomatic infection also leads to immunity, and therefore local depletion of susceptible individuals around infectious individuals. Hence, for the same relative infectiousness, the contribution of asymptomatic individuals to transmission is much lower when spatial heterogeneity is taken into account.

Nonetheless, our results suggest that asymptomatic individuals do contribute a small amount to transmission and that they can “bridge” gaps between VL cases in transmission chains, as the best-fitting model has non-zero asymptomatic relative infectiousness. Superficially, this appears to conflict with preliminary results of xenodiagnosis studies in which asymptomatic individuals have failed to infect sandflies according to microscopy (44). However, historical (12, 45) and experimental (46) data show that provision of a second blood meal and optimal timing of sand fly examination are critical to maximizing sensitivity of xenodiagnosis. These data suggest that recent xenodiagnosis studies (11, 44), in which dissection occurred within 5 days of a single blood meal, may underestimate the potential infectiousness of symptomatic and asymptomatic infected individuals. Occurrence of VL in isolated regions where there are asymptomatically infected individuals, but virtually no reported VL cases (27, 47), also seems to suggest that asymptomatic individuals can generate VL cases. However, it is possible that some individuals who developed VL during the study went undiagnosed and untreated, and that we have inferred transmissions from asymptomatic individuals in locations where cases were missed. We will investigate the potential role of under-reporting in future work.

The analysis presented here is not without limitations. As can be seen from the model simulations (Fig. S20), the model is not able to capture the full spatiotemporal heterogeneity in the observed VL incidence when fitted to the data from the whole study area, as it underestimates the number of cases in higher-incidence paras (e.g. paras 1, 4 and 12). There are various possible reasons why the incidence in these paras might have been higher, including higher sandfly density, lower initial levels of immunity, variation in infectiousness between cases and within individuals over time, dose-dependence in transmission (whereby flies infected by VL cases are more likely to create VL cases than flies infected by asymptomatic individuals (22)), and variation in other unobserved risk factors (such as bed net use). It was not possible to include sandflies explicitly in the model due to an absence of data on sandfly abundance and gaps in understanding of *P. argentipes* bionomics (48). We were unable to incorporate variation in infectiousness between individuals in the same disease state and over time within disease states due to the relatively limited xenodiagnostic data available on infectiousness of VL and PKDL and lack of data on variation in infectiousness of individuals over time (e.g. from serial parasite load measurements or serial xenodiagnosis). We were also not able to consider the role of HIV-VL co-infected individuals in transmission as there was no data on HIV infection in the study population, but other data suggests they may contribute significantly with prevalences of HIV coinfection of up to 6% in India (49) and higher infectiousness towards sandflies (50). Further laboratory and field studies are needed to quantify these sources of heterogeneity to be able to parameterise variation in transmission intensity between locations.

Nevertheless, our analysis provides unique insights into how visceral leishmaniasis spreads in space and time and the role played by PKDL and asymptomatic infection in this process. We have developed a novel MCMC data augmentation framework to account for the endemic nature of the disease and high proportion of asymptomatic infection, and used it to generate quantitative estimates for guiding targeted interventions around VL and PKDL cases. In future work we will predict the impact of different spatiotemporally-targeted interventions on VL incidence using the simulation model developed here.

## Materials and Methods

### Data Collection

The data used in this study was collected in 4 household surveys conducted between July 2007 and December 2010 in a highly VL-endemic community in Fulbaria upazila, Mymensingh district, Bangladesh (for full details of the study protocol and case definitions see (8)). The GPS coordinates of all households were recorded using a Garmin 76 GPS receiver, and the migration of individuals within, and in and out of, the study area was recorded.

### Transmission Model

We developed a discrete-time individual-level spatial kernel transmission model for VL by extending our previous individual-level model (51) to explicitly include asymptomatic infection and PKDL. In the model, the infection pressure on susceptible individual *i* in month *t* is given by the sum of the individual infection pressures on them from surrounding infectious individuals (*j ϵ Inf* (*t*)), which are a function of their distance *d*_*ij*_ from *i* and their relative infectiousness (compared to VL cases) *h*_*j*_ (*t*), plus a background transmission rate *E* to account for unexplained infections:

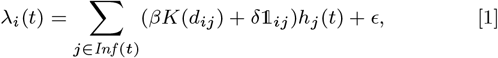

where *K*(*d*) = *e*^*−d/α*^ is the spatial kernel function that determines how transmission risk decreases with distance (with distance decay rate 1*/α*), *β* is the spatial transmission rate constant, *δ* is the extra within-household transmission rate, and 𝟙_*ij*_ is an indicator function that is 1 if *i* and *j* share the same household and 0 otherwise. A proportion *p*_*I*_ of infections lead to VL following a negative-binomially-distributed NB(*r, p*) incubation period, while the remaining infections are asymptomatic with geometric Geom(*p*_2_) duration. We use *p*_*I*_ = 0.15, *r* = 3 and *p*_2_ = 1*/*5 based on previous analyses (30, 51), and estimate *p* along with the transmission parameters.

We assume individuals’ relative infectiousnesses *h*_*j*_ (*t*) remain constant in each infection state and parameterise those of VL and PKDL cases using data from a recent xenodiagnosis study in Bangladesh (11), and those of asymptomatic and pre-symptomatic individuals based on estimates from previous modelling studies (23, 43) and preliminary evidence from a xenodiagnosis study in India that nonsymptomatic infected individuals are much less infectious than VL or PKDL cases (44). Given the uncertainty in the infectiousness of asymptomatic and pre-symptomatic individuals and the absence of experimental data on their relative infectiousness, we assume they are equally infectious and compare the fit of the model with different fixed values for their relative infectiousness from 0 to 2% using the Deviance Information Criterion. We also compare the fit of models without and with additional within-household transmission (*δ* = 0 vs *δ >* 0).

### Bayesian Data Augmentation

We estimated the parameters in the transmission model, ***θ*** = (*β, α, ϵ, δ, p*), the unobserved infection times of VL cases and infection and recovery times of asymptomatic individuals, and individuals’ unobserved initial statuses by sampling from the joint posterior distribution of ***θ*** and the missing data **X** given the observed data **Y** (months of birth, migration, and death; VL and PKDL onset and recovery times; etc.), ℙ(***θ*, X**|**Y**) *∝ L*(***θ***; **Y, X**) ℙ (***θ***), where *L*(***θ***; **Y, X**) denotes the complete data likelihood and ℙ (***θ***) is the prior distribution for ***θ***, using a Bayesian data augmentation framework (see *SI Text* for full details). Markov chain Monte Carlo (MCMC) methods were used to obtain the joint posterior distribution by iteratively sampling from the posterior distribution of the parameters given the observed data and current value of the missing data, ℙ (***θ***|**Y, X**), and the posterior distribution of the missing data given the observed data and the current values of the parameters, ℙ (**X**|**Y, *θ***) (52). Relatively uninformative Gamma distributions were used for the priors for the transmission parameters (*β, α, ϵ* and *δ*), and a relatively informative conjugate Beta prior was used for the incubation period distribution parameter *p* based on a previous estimate of the mean incubation period and its uncertainty (30) (see *SI Text* for further details).

Once the posterior distribution of the parameters and missing data had been obtained from the MCMC, 1,000 samples were drawn from the posterior distribution and the posterior predictive distributions of infection sources for all infectees derived for each sample. These were used to draw an infector for each infectee to reconstruct the transmission tree. Thus we obtained a set of 1,000 possible transmission trees that accounted for uncertainty in the parameter values, infection times, infection sources and individuals’ initial statuses. The mean distance from each infector to their infectees and time from their onset to the infections of their infectees was calculated for each tree, and then averaged over all trees in which that individual was an infector to obtain distributions of mean distances and times to infectees across all infectors (Figures 4A and 4B). The posterior predictive distributions of infection sources were also used to estimate the number of secondary infections for each asymptomatic individual, VL case and PKDL case (Figures 4C, S18 and S19B), and the time-dependent effective reproduction number (Fig. S19C).

### Model Simulations

We implemented a simulation version of the transmission model (full details in *SI Text*) to assess the ability of the model to reproduce the observed data and to investigate the counterfactual impact of different hypothetical interventions against PKDL on VL incidence. One hundred samples of the parameters and individuals’ infection statuses in December 2002 were drawn from the posterior distribution obtained from the MCMC and 100 simulations of the model run for each sample starting from January 2003 (at which point all but one of the paras had had at least 1 VL case since January 2002), to give 10,000 realisations of the epidemic under “normal” interventions. This process was then repeated with PKDL infectiousness set to zero (to simulate no development of PKDL), and then again with the mean duration of PKDL infectiousness halved (to simulate more rapid detection and treatment of PKDL), and the percentage difference in the total number of cases in each “alternative-intervention” simulation from that in each “normal-intervention” simulation calculated.

### Ethical Approval

The study was approved by the institutional review boards of the International Centre for Diarrhoeal Disease Research, Bangladesh (protocol #2007-003) and the Centers for Disease Control and Prevention (protocol #5065), and informed consent was obtained from all participants or parents/guardians in the case of children. The analysed data contains personally identifiable information and so cannot be made freely available. Individuals who wish to access the data should contact.

## Data Availability

The data analysed in this study contains personally identifiable information about a stigmatising disease and so cannot be made freely available. Individuals who wish to access the data should contact. All code used in the study is freely available from https://github.com/LloydChapman/VLSpatiotemporalModelling.

## ACKNOWLEDGMENTS

This work was supported by the Bill and Melinda Gates Foundation through the SPEAK India consortium (Grant OPP1183986) (L.A.C.C., M.M.C., G.F.M.) and the NTD Modelling Consortium (Grant OPP1184344) (L.A.C.C., S.E.F.S., T.M.P., T.D.H., G.F.M.). S.E.F.S. gratefully acknowledges funding from MRC (Grant MR/P026400/1) and EPSRC (Grant EP/R018561/1). L.A.C.C. wishes to thank Marinella Capriati, Jim and Monica Chapman and members of the Centre of Mathematical Modelling of Infectious Diseases, London School of Hygiene and Tropical Medicine for very useful discussions.

## Supporting Information

This file provides further information on the spatiotemporal transmission model and Bayesian inference framework described in the *Materials and Methods* section in the main text, including a full description of the model, the formula for the likelihood of the model, details of the Markov Chain Monte Carlo (MCMC) algorithm, and additional output from the MCMC algorithm.

### Model description

The model used in this paper (Fig. S1) is an extension of our previously published individual-level spatiotemporal SEIR model of visceral leishmaniasis (VL) transmission (1) to explicitly include asymptomatic infection and post-kala-azar dermal leishmaniasis (PKDL), and account for unobserved initial infection statuses and migration of individuals. We measure time in units of months, with *t* = 1 corresponding to the start of the study (January 2002) and *t* = 108 the end of the study (December 2010), and label individuals who developed VL symptoms between January 2002 and December 2010 by *i* = 1, 2,*…, n*_*I*_ (*n*_*I*_ = 1018) and the remainder of the population by *i* = *n*_*I*_ + 1, *n*_*I*_ + 2,*…,n* (*n* = 25, 506). Between January 2002 and December 2010, 725 individuals relocated between different households within the study area, which we accounted for by including a second observation for these individuals in their second household. These internal migrators were essentially treated as extra individuals for the purpose of calculating pairwise distances and transmission rates. However, their infection status was updated in such a way that it was consistent across the two observations (e.g. if they were asymptomatically infected in the period of the first observation and did not recover before relocating, their asymptomatic infection status was carried over to the second observation). We denote the vectors of times of birth, immigration, emigration, asymptomatic infection, pre-symptomatic infection, VL onset, recovery from VL through treatment or naturally from asymptomatic infection, VL relapse, VL relapse treatment, temporary recovery to dormant infection prior to PKDL, PKDL onset, PKDL resolution and death for all individuals by **B** = (*B*_*i*_), **IM** = (*IM*_*i*_), **EM** = (*EM*_*i*_), **A** = (*A*_*i*_), **E** = (*E*_*i*_), **I** = (*I*_*i*_), **R** = (*R*_*i*_), **I**_*R*_ = (*I*_*Ri*_), **R**_*R*_ = (*R*_*Ri*_), **D** = (*D*_*i*_), **P** = (*P*_*i*_), **R**_*P*_ = (*R*_*Pi*_), and **M** = (*M*_*i*_) (*i* = 1,*…, n*) respectively. In any particular month *t* individuals can be in one of the 7 states shown in Figure S1A:

- susceptible: *S*(*t*) := *{i* : max(*B*_*i*_, *IM*_*i*_) *≤ t<* min(*EM*_*i*_, *A*_*i*_, *E*_*i*_, *M*_*i*_)*}*
- asymptomatically infected (and potentially infectious): *𝒜*(*t*) := *{i* : *A*_*i*_ *≤ t<* min(*EM*_*i*_, *R*_*i*_, *D*_*i*_, *M*_*i*_)*}*
- pre-symptomatically infected (and potentially infectious): *ϵ* (*t*) := *{i* : *E*_*i*_ *≤ t< I*_*i*_*}*
- symptomatic VL: *ℐ*(*t*) := *{i* : max(*I*_*i*_, *IM*_*i*_) *≤ t<* min(*EM*_*i*_, *R*_*i*_, *D*_*i*_, *M*_*i*_) or *I*_*Ri*_ *≤ t<* min(*R*_*Ri*_, *D*_*i*_, *M*_*i*_)*}*
- dormantly infected (i.e. treated for VL or recovered from asymptomatic infection but with subsequent VL relapse or PKDL): *𝒟*(*t*) := *{i* : *D*_*i*_ *≤ t<* min(*I*_*Ri*_, *P*_*i*_)*}*
- symptomatic PKDL (i.e. visible skin lesions): 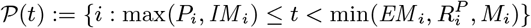
- recovered (i.e. treated for primary VL, VL relapse or PKDL, or self-resolved from PKDL, or recovered from asymptomatic infection): 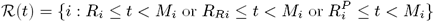.

Upon infection, individuals either develop pre-symptomatic infection with probability *p*_*I*_ or asymptomatic infection with probability 1 *− p*_*I*_ (see Table S2 for values of fixed parameters used in the model). Pre-symptomatic individuals progress to symptomatic VL following a variable incubation period, and then to recovery, or dormant infection if they later relapse or develop PKDL, upon treatment. VL cases that relapse either return to dormant infection or progress to recovery once re-treated. Asymptomatically infected individuals either recover naturally or very occasionally progress to dormant infection and subsequent PKDL. PKDL cases can either resolve following treatment or naturally, whereupon they enter the recovered class. We assume that recovered individuals do not return to being susceptible, i.e. cannot be reinfected, irrespective of whether they have recovered from VL, PKDL or asymptomatic infection. It is still uncertain whether individuals can become reinfected with *L. donovani* parasites, particularly asymptomatically infected individuals. Most previous modelling studies have assumed or concluded that individuals can be reinfected (2–4), but have not accounted for spatial variation in transmission and available evidence suggests that repeat episodes of VL are relatively rare and are due to relapse not reinfection (5–7).

For each individual *i* in the study population, we define *V*_*i*_ = max(0, *B*_*i*_, *IM*_*i*_) and *W*_*i*_ = min(*T* + 1, *EM*_*i*_, *M*_*i*_) as their times of entry into and exit from the study area respectively. All non-symptomatic individuals (individuals without any VL or PKDL symptoms before or during the study) who were born or entered the study area after January 2002 are assumed to have been susceptible upon entry.

#### Pairwise transmission rates

Susceptible individuals become infected either from pre-symptomatically infected individuals, from symptomatic VL cases, PKDL cases, asymptomatic individuals, or ‘background’ (unexplained) transmission. The transmission rate, or ‘infection pressure’, between an infected individual *j* and a susceptible individual *i* at time *t* is given by:

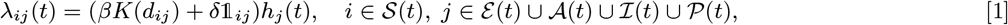

where *β* is the rate constant for spatial transmission between infected and susceptible individuals; *K*(*d*_*ij*_) is the spatial kernel function that scales the transmission rate by the distance *d*_*ij*_ between individuals *i* and *j*; *δ* (*≥*0) is a rate constant for additional within-household transmission; 𝟙_*ij*_ is an indicator function for individuals living in the same household, i.e.

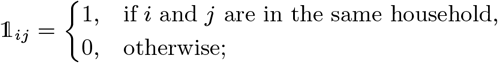

and *h*_*j*_ (*t*) is the infectiousness of *j* at time *t*. Individuals are assumed to be stationary in their households when transmission to and from sandflies occurs (since the vast majority of sandfly biting occurs at night when individuals are asleep in, or directly outside, their homes (8–13)), so that the distances between them *d*_*ij*_ are fixed when transmission takes place. However, the changes in the distances between individuals that occur when individuals migrate is incorporated into the model (by calculating the distances between migrators and all other individuals both when they are in their first household and when they are in their second household).

**Fig. S1.**
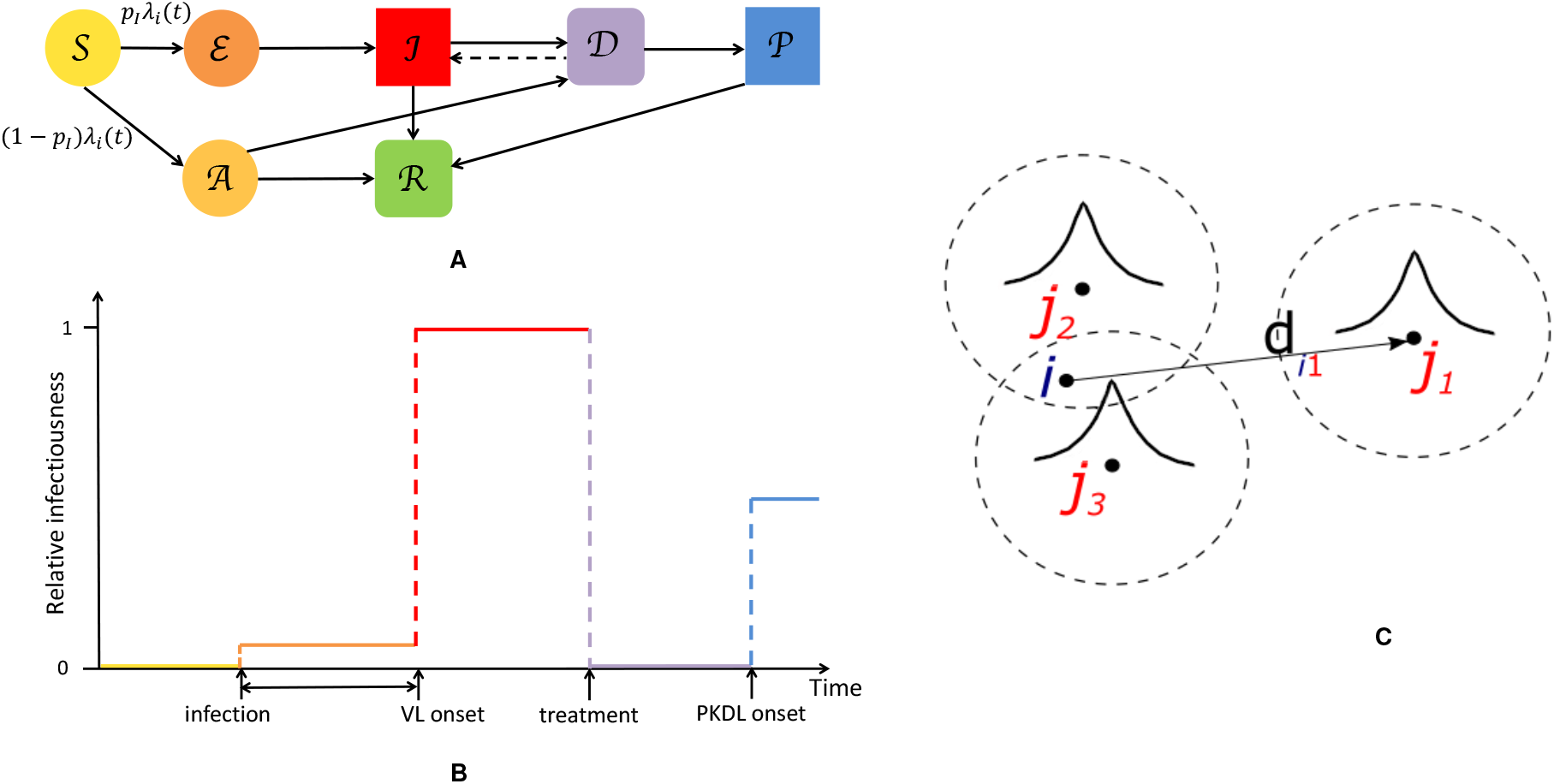
Individual-level spatiotemporal transmission model. (A) States in the model (*S* = susceptible, *𝒜* = asymptomatically infected, *ε* = pre-symptomatically infected, *ℐ* = symptomatic VL, *𝒟* = dormantly infected, *𝒫* = PKDL and *ℛ* = recovered). Squares represent fully observed states, rounded-edge squares partially observed states, and circles unobserved states. *p*_*I*_ = probability of symptomatic infection, *λ*_*i*_ *t* = total infection pressure on susceptible individual *i* at time *t*. (B) Relative infectiousness over time of individual passing through top pathway (symptomatic infection) in (A). Diagram not to scale and shown assuming that pre-symptomatic individuals are infectious and VL case developed macular PKDL. (C) The total infection pressure *λ*_*i*_(*t*) on a susceptible individual *i* at time *t* is the sum of the individual infection pressures on them from infectious individuals around them (here *j*_1_, *j*_2_ and *j*_3_), which are a function of how far away they are (depicted by the curves), the time since their infections and whether their infection is symptomatic or asymptomatic. *d*_*i*1_ = distance between *i* and *j*_1_.

Since we found little difference in the goodness of fit of the model (based on the Deviance Information Criterion (DIC)) between an exponentially decaying spatial kernel and a Cauchy-type kernel in our previous study (1) (the exponential kernel gave a marginally better fit), we use the exponential kernel here:

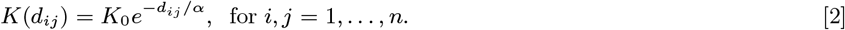

where 1*/α* is the distance decay rate (per m) in transmission risk (so smaller values of *α* correspond to a faster decrease in risk with distance), and *K*_0_ is a normalisation constant, defined such that

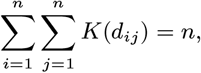

which reduces correlation between *α* and *β* and thus improves the mixing of the MCMC chain. However, unlike in our previous study, we allow the possibility of transmission between individuals in different paras (hamlets). The reason for this is that surveying of neighbouring paras was more complete in the two clusters of paras in the present study, and thus some minimum inter-para household distances were less than intra-para household distances and within the maximum reported flight range of *P. argentipes* sandflies (a few hundred metres (14–17)), so transmission between neighbouring surveyed paras may have occurred.

#### Infectiousness over time

There is relatively little data available on the infectiousness of individuals in different *L. donovani* infection states over time. We therefore assume that the infectiousness of individuals remains constant within each state and individuals in the same state have the same probability of passing on infection, such that an individual’s infectiousness over time takes the form of a step function (Fig. S1B). The relative infectiousness of asymptomatic individuals compared to VL cases, *h*_4_, is assumed to take a fixed value between 0 and 0.02 based on estimates from previous modelling studies (3, 4, 18) and preliminary results from xenodiagnosis studies that have so far not found infection in flies fed on asymptomatically infected individuals (19). We test different values of *h*_4_ to assess whether asymptomatic individuals are in fact infectious towards sandflies (see *Model Comparison* below). In the absence of data on the relative infectiousness of pre-symptomatic individuals and asymptomatic individuals, we take the relative infectiousness of pre-symptomatic individuals, *h*_0_, to be the same as that of asymptomatic individuals (i.e. *h*_0_ = *h*_4_).

One-hundred and thirty-eight of the 190 PKDL cases underwent one or more examinations by a trained physician to determine the type and extent of their lesions (Table S1). Data from a recent xenodiagnosis study in Bangladesh (20) on 47 PKDL patients (21 nodular and 26 macular/papular) and 15 VL patients is used to assign infectiousness to these cases according to their lesion type (macular/papular, plaque, or nodular) (Table S1). The results of the xenodiagnosis study suggest that infectiousness of PKDL increases with lesion severity and macular/papular PKDL cases are less infectious towards sandflies than VL cases but nodular PKDL cases more so. As plaques were not treated as a separate lesion type in the study, but are intermediate in severity between macular/papular lesions and nodular lesions (21), a value halfway between the infectiousness of macular/papular PKDL cases and nodular PKDL cases is assigned to these individuals. The 52 PKDL cases that were not physically examined during the study are assigned an average of the infectiousnesses of the different lesion types (weighted by frequency amongst the examined cases). Although it is not known for certain whether treated VL cases who subsequently relapse or develop PKDL are uninfectious following treatment, we assume dormantly infected individuals are uninfectious, based on rapid decreases in parasitaemia observed in VL cases following the start of treatment (22, 23)). We assume that relapse cases are as infectious upon relapse as in their first clinical episode, as relapse appears to be associated with resurgence in parasitaemia to high parasite loads (23, 24). Over half (100/190=53%) of the PKDL cases did not receive treatment, and of these 49 self-resolved in a median time of 20 months (interquartile range (IQR) 14–32 months). These cases are assumed to have remained infectious until their lesions resolved, while treated PKDL cases are assumed to have stopped being infectious once their treatment commenced (25).

**Table S1.** Infectiousnesses of different infection states relative to visceral leishmaniasis (VL).

| Infection state | Proportion in physical examinations | Proportion positive in xenodiagnosis experiments* | Relative infectiousness | References |
| --- | --- | --- | --- | --- |
| Pre-symptomatic | - | - | $h_0 = 0-0.02$ | Assumed ($h_0 = h_4$) |
| VL (primary/relapse) | - | 10/15 (67%) | 1 | Reference value |
| PKDL |  |  |  |  |
| -macular/papular | 101/138 (73%) | 9/26 (35%) | $h_1 = (9/26)/(10/15) = 0.52$ | (20, 21) |
| -plaque | 31/138 (22%) | - | $h_2 = ((9/26 + 18/21)/2)/(10/15) = 0.90$ | (21) |
| -nodular | 6/138 (4%) | 18/21 (86%) | $h_3(18/21)/(10/15) = 1.29$ | (20, 21) |
| -unexamined | - | - | $h_u = 101/138 \times 0.52 + 31/138 \times 0.90 + 6/138 \times 1.29 = 0.64$ | Assumed |
| Asymptomatic | - | - | $h_4 = 0-0.02$ | Assumed from (4, 18) |

\* Proportion who infected flies based on positive microscopy and/or qPCR on fed flies.

Thus, individual *j* ‘s infectiousness at time *t* is given by

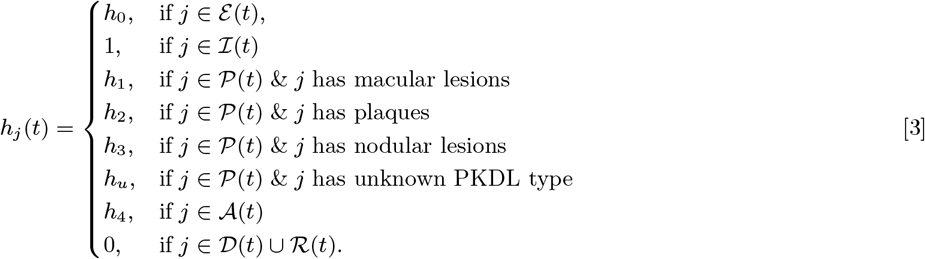

#### Incubation period

Following previous work (1), we model the incubation period as negative binomially distributed NB(*r, p*) with fixed shape parameter *r* = 3 and ‘success’ probability parameter *p*, and support starting at 1 (such that the minimum incubation period is 1 month):

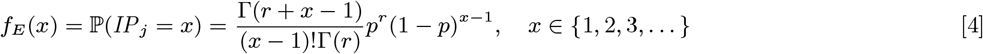

We estimate *p* in the MCMC algorithm for inferring the model parameters and missing data (see below).

#### VL onset-to-treatment time distribution

Several VL cases with onset before 2002 have missing symptom onset and/or treatment times (only their onset year is recorded), and may therefore have been infectious at the start of the study period. In order to be able to infer the onset-to-treatment times of these cases, 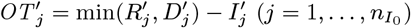, in the MCMC algorithm (see below) we model the onset-to-treatment time distribution as a negative binomial distribution NB(*r*_1_, *p*_1_) and fit to the onset-to-treatment times of all VL cases for whom both onset and treatment times were recorded (Figure S2A):

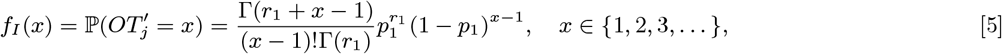

to obtain *r*_1_ = 1.34 and *p*_1_ = 0.38 (corresponding to a mean onset-to-treatment time of 3.2 months).

#### Asymptomatic infection duration

Neither the infection or recovery time of asymptomatically infected individuals is observed, so we need to specify a distribution for the asymptomatic infection duration in order to infer their recovery times. Based on a previous multi-state Markov model of the natural history of VL (26), in which it was assumed that the duration of asymptomatic infection is exponentially distributed and its mean was estimated as approximately 5 months, we assume the asymptomatic infection duration, *AIP*_*j*_ = *R*_*j*_ *− A*_*j*_, follows a geometric distribution (the discrete analog of the exponential distribution) Geom(*p*_2_), with a minimum duration of 1 month and a mean of 5 months 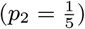:

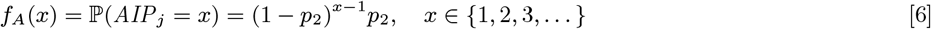

The choice of the geometric distribution is partly motivated by the fact that it is a memoryless distribution, i.e. P(*AIP*_*j*_ *> s* + *t* |*AIP*_*j*_ *> s*) = ℙ(*AIP*_*j*_ *> t*). This simplifies the estimation of recovery times for initially asymptomatically infected individuals (see *Model for initial status of non-symptomatic individuals* below) in the MCMC algorithm as it means that the probability that an initially asymptomatically infected individual remains infected for a further *t* months is the same as if they were infected in month 0 regardless of when they were infected.

#### Dormant infection duration

Sixteen (8.4%) of the 190 PKDL cases reported no prior history of VL, and are assumed to have been previously asymptomatically infected. We assume that their duration of dormant infection prior to PKDL and that of VL cases who develop PKDL follow the same negative binomial distribution, NB(*r*_3_, *p*_3_), and estimate *r*_3_ and *p*_3_ by fitting to the observed VL-treatment-to-PKDL-onset times by maximum likelihood estimation (Figure S2B). Unlike for the incubation period, we take 0 months to be the minimum duration of dormant infection, since two VL cases developed PKDL in the same month as they were treated for VL, and one had simultaneous VL and PKDL. Thus the probability mass function (PMF) is:

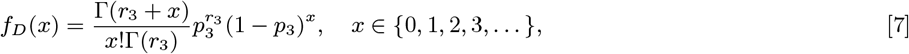

where *r*_3_ = 1.73 and *p*_3_ = 0.065 (corresponding to a mean duration of dormant infection of 25 months).

**Fig. S2.**
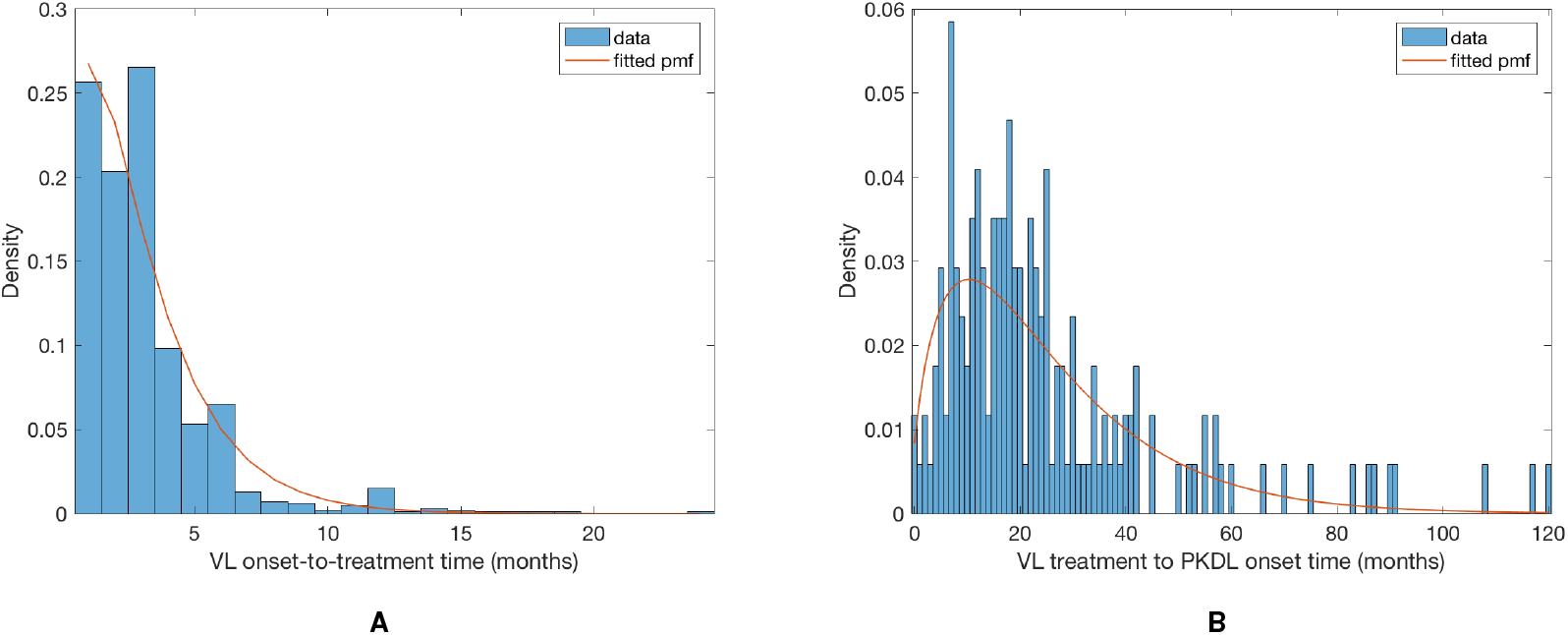
Observed and fitted negative binomial distributions of (A) VL onset-to-treatment time and (B) VL-treatment-to-PKDL-onset time.

#### Time to relapse and relapse duration

Forty-five VL cases suffered treatment failure or relapse during the study. Of these, 16 were treated with miltefosine and 29 with sodium stiboglucanate (SSG). Of those treated with miltefosine, 15 were treated with counterfeit drug in 2008 (27) and all but one of these cases reported no gap between the start of treatment and recurrence of symptoms (the other case reported a gap of 30 days). We assume that individuals without any gap between treatment and symptom recurrence suffered treatment failures, and that their infectiousness continued for 1 month following the start of treatment until they were treated with SSG. The gap between the start of treatment and new symptoms occurring was recorded for 7 of the cases originally treated with SSG or miltefosine from a clinical trial, and was non-zero in all cases. We assume that all cases not recorded as having immediate recurrence of symptoms suffered treatment relapse and that the time to relapse follows a geometric distribution Geom(*p*_4_) with PMF:

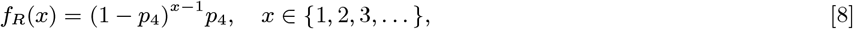

where fitting to the recorded gaps gives *p*_4_ = 0.13 (corresponding to a mean time to relapse of 7.9 months). Relapse cases are assumed to be uninfectious from their treatment month to their relapse time and their duration of symptoms upon relapse is assumed to follow the same distribution as the onset-to-treatment time for a first VL episode (Eq. (5)). We assume all relapse cases were treated for relapse before the end of the study, since the latest treatment time for primary VL in a case that subsequently relapsed was April 2009.

#### Infection pressure

The total infection pressure on susceptible individual *i* at time *t* is given by the sum of the infection pressures on them from all infectious individuals (VL cases, PKDL cases, pre-symptomatic individuals and asymptomatic individuals) at time *t* in Eq. (1) (see Fig S1C) plus a constant background transmission rate *ϵ* to account for unexplained infections due to non-explicitly modelled factors (e.g. due to short-term movement of individuals)

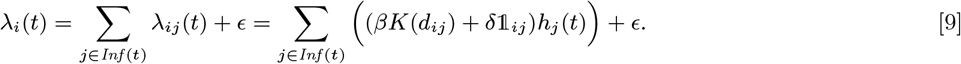

where *Inf* (*t*) = *ε* (*t*) *∪𝒜* (*t*) *∪ℐ* (*t*) *∪𝒫* (*t*) is the set of all individuals infectious at time *t*. The transmission process is thus described by a discrete-time approximation to an inhomogeneous Poisson process with rate *λ*(*t*) =∑_*i∈S*(*t*)_ *λ*_*i*_(*t*). The probability of susceptible individual *i* remaining susceptible in any particular month *t* is:

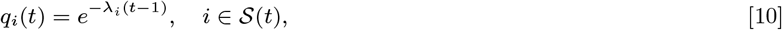

while the probabilities of pre-symptomatic or asymptomatic infection in month *t* given susceptibility up to month *t −* 1 are, respectively:

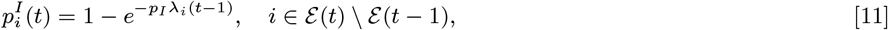

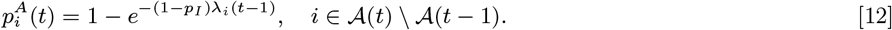

### Model for initial status of non-symptomatic individuals

As there was transmission and VL in the population before the start of the study, individuals with no record of VL prior to 2002 may have been asymptomatically infected before the start of the study, i.e. the initial statuses of non-symptomatic individuals are censored and need to be estimated. Although the data on VL pre-2002 is likely incomplete, we adopt the simplifying assumption that any individual that had VL prior to 2002 is at least recorded as having had previous VL (even if their onset and treatment times are missing, imprecise or inaccurate), such that anyone in the rest of the population who was infected prior to 2002 could only have had asymptomatic infection. We also average over historical and spatial variation in the transmission rate, by assuming that the asymptomatic infection rate prior to 2002, *λ*_0_, was constant. We assume that all 16 individuals who developed PKDL during the study period without prior VL were asymptomatically infected during the study rather than before (i.e. we ignore the possibility that such individuals were initially dormantly infected). The latter assumption is not unreasonable despite the estimated long duration of dormant infection, as the earliest PKDL onset amongst these individuals was in November 2005 (47 months into the study), and it is unlikely to significantly affect the results given the small number of such cases. With these assumptions we arrive at the simplified model for the initial status of non-symptomatic individuals shown in Figure S3.

**Fig. S3.**
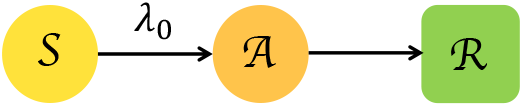
Model for the initial statuses of non-symptomatic individuals, with a constant asymptomatic infection rate, *λ*_0_.

The probabilities of each non-symptomatic individual initially present (i.e. with *V*_*j*_ = 0) being susceptible, asymptomatically infected, or recovered from asymptomatic infection at time *t* = 0 can then be found by calculating the probability of avoiding infection in every month from their birth to the start of the study, summing over the probabilities of being infected in one of the months between their birth and the start of the study and recovering after the start of the study, and summing over the probabilities of being infected in a month before the start of the study and recovering before the start of the study, respectively:

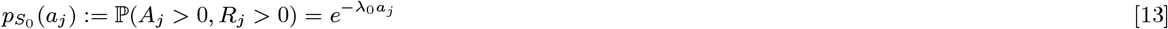

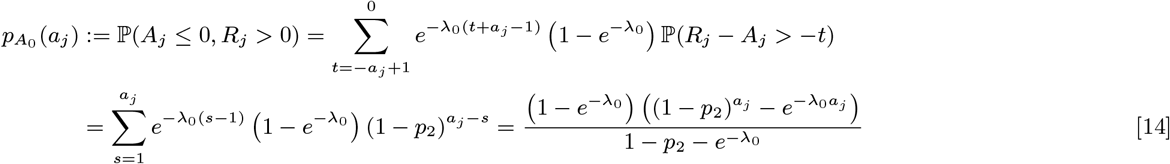

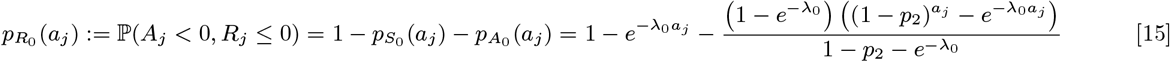

where *a*_*j*_ is the age of individual *j* in months at *t* = 0. Since we assume that non-symptomatic individuals who are born, or who immigrate into the study area, after the start of the study (with *V*_*j*_ *>* 0) are susceptible, for notational convenience we define the probabilities for these individuals as 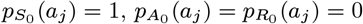.

**Fig. S4.**
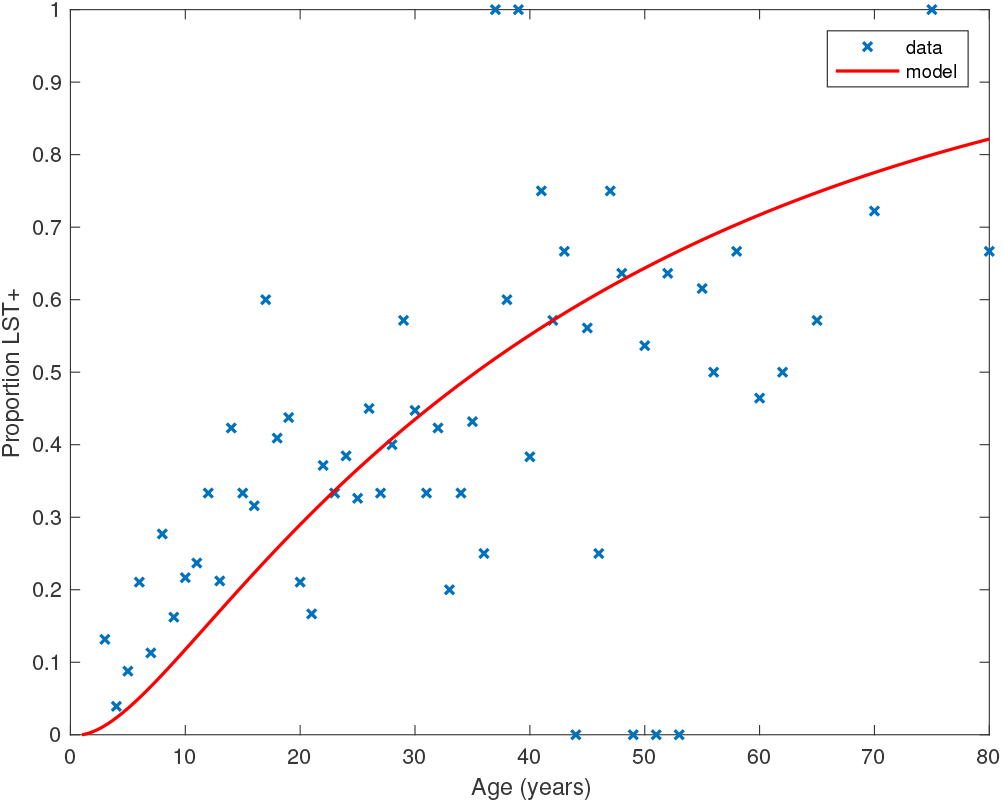
Age-prevalence distribution of LST positivity among non-symptomatic individuals in three of the study paras in 2002 and fit of initial status model in Figure S3.

We estimate the historical asymptomatic infection rate, *λ*_0_, by fitting the model to age-prevalence data on leishmanin skin test (LST) positivity amongst non-symptomatic individuals from a cross-sectional survey of three of the study paras conducted in 2002 (28) (see Figure S4). We assume that entering state *ℛ* corresponds to becoming LST-positive, as LST positivity is a marker for durable, protective cell-mediated immunity against VL (28, 29), and estimate *λ*_0_ by maximising the binomial likelihood:

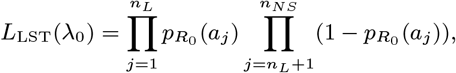

where *n*_*L*_ = 479 is the number of non-symptomatic individuals that were LST-positive out of *n*_*NS*_ = 1399 individuals tested. This gives *λ*_0_ = 0.0019 mnth^*−*1^ (95% CI 0.0017–0.0021 mnth^*−*1^).

### Complete data likelihood

We assume that the end of the epidemic was observed from the point of view of VL and PKDL cases, i.e. every individual in the study population who eventually developed VL or PKDL did so within the observation period. Whilst it is likely that there was ongoing transmission beyond the end of the study in December 2010, this simplifying assumption should not introduce significant error based on the epidemic curves in Figure 1 in the main text and the very low numbers of VL and PKDL cases with onset in the final months of the study.

As noted in the main text, there is a considerable amount of missing data, including the infection times of VL cases (**E**) and the infection and recovery times of asymptomatic individuals (**A** and **R**_*A*_). So that we can specify the complete data likelihood — the likelihood of all events if all variables had been observed — and perform likelihood-based inference, we define the sets of all observed data and missing data as 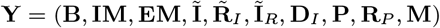 and 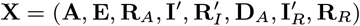 respectively, where 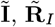 and 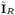 are the observed onset and recovery times of VL cases and relapse times of VL relapse cases; 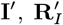 and 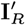 are their missing counterparts; and **D**_*I*_ and **D**_*A*_ are the start times of dormant infection for VL cases and asymptomatic individuals.

**Table S2.** Values of fixed parameters used in the model

| Parameter | Description | Value | Source |
| --- | --- | --- | --- |
| $p_I$ | Proportion of infected individuals who develop VL | 0.15 | (26) |
| $r$ | Shape parameter of negative binomial incubation period distribution | 3 | (1) |
| $r_1$ | Shape parameter of negative binomial onset-to-treatment time distribution | 1.34 | Fitting to observed data |
| $p_1$ | 'Success' probability parameter of negative binomial onset-to-treatment time distribution | 0.38 | Fitting to observed data |
| $p_2$ | Asymptomatic infection duration distribution parameter | 1/5 | (26) |
| $r_3$ | Shape parameter for negative binomial dormant infection duration distribution | 1.77 | Fitting to observed data |
| $p_3$ | 'Success' probability parameter of negative binomial dormant infection duration distribution | 0.066 | Fitting to observed data |
| $p_4$ | Treatment-to-relapse distribution parameter | 0.13 | Fitting to observed data |
| $\lambda_0$ | Historical asymptomatic infection rate | 0.0019 | Fitting to data on LST-positive prevalence in study area (28) |
| $r_5$ | Shape parameter of negative binomial PKDL-onset-to-recovery time distribution* | 1.18 | Fitting to observed data |
| $p_5$ | 'Success' probability parameter of negative binomial PKDL-onset-to-recovery time distribution* | 0.066 | Fitting to observed data |
| $p_P$ | Proportion of VL cases who develop PKDL* | 0.17 | Survival analysis in (21) |
| $p_D$ | Proportion of asymptomatic individuals who develop PKDL* | 0.0028 | Estimated from observed incidence of PKDL amongst asymptomatic individuals |

\*Parameters only used in the simulations in *Model simulations* as they do not appear in the likelihood in Eq. (16).

With these definitions, the complete data likelihood for the augmented data **Z** = (**Y, X**) given the model parameters ***θ*** = (*β, α, ϵ, δ, p*) is composed of the products of the probabilities of all the different individual-level events over all months:

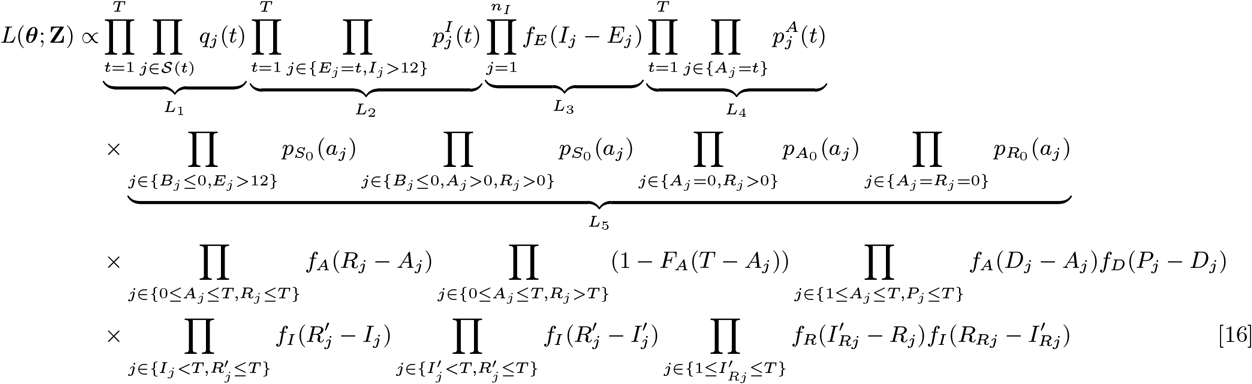

where *F*_*A*_(*x*) = 1 *−* (1 *− p*_2_)^*x*^ (*x ≥* 1) is the cumulative distribution function of the asymptomatic infection period distribution (Eq. (6)).

Given the length and variability of the incubation period, some VL cases with onset early in the study (i.e. in 2002) may have been infected before 2002. However, unlike in our previous work (1), we cannot assume that these cases arose due to background transmission, as there was not an extended period without VL cases directly before 2002. In fact, 413 individuals in the study area were recorded as having VL with onset prior to 2002, 141 of these in 2000 or 2001. Since the data on VL occurrence, and onset and treatment times before 2002 is less complete and probably less reliable than from 2002 onwards, there may be missing information on potential sources of infection of cases with onset during 2002. We therefore exclude these cases from the infection probability part of the likelihood (*L*_2_), but still impute their infection times in the MCMC algorithm (see below) by drawing new infection times for them using the incubation period distribution, and accepting/rejecting these based on the effect they have on the likelihood of the infection times of other individuals.

## Parameter estimation

### Bayesian inference

Since the infection times of VL cases (**E**) and infection and recovery times of asymptomatic individuals (**A** and **R**_*A*_) were unobserved, and some recovery, relapse and relapse treatment times of VL cases 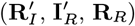 were not recorded, and it is not computationally feasible to sum over all possible configurations of this missing data to calculate the complete data likelihood (Eq. (16)), we treat the missing times as extra parameters and use a data augmentation approach to sample from the joint posterior distribution of the model parameters ***θ*** = (*β, α, ϵ, δ, p*) and the missing data **X** given the observed data **Y**

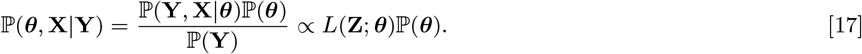

We do this using a Metropolis-within-Gibbs MCMC data augmentation algorithm in which we iterate between sampling from the conditional posterior distribution of the parameters given the observed data and the current value of the missing data, ℙ (***θ***|**Y, X**), and the conditional posterior distribution of the missing data given the current parameter values and the observed data, ℙ (**X**|***θ*, Y**) (1, 30).

### MCMC data augmentation scheme

#### Asymptomatic infection times

In order to account for the uncertainty in the parameter estimates due to the missing asymptomatic infection and recovery times, we need to estimate which individuals were asymptomatically infected during the study and when they were infected, as part of the MCMC algorithm. To do this, we assign every non-symptomatic individual *j* in the population an asymptomatic infection time and recovery time pair, (*A*_*j*_, *R*_*j*_) *∈* {*V*_*j*_, *V*_*j*_ + 1,*…, W*_*j*_ *−* 1,*T* + 1} *×*{*A*_*j*_ + 1, *A*_*j*_ + 2,*…, W*_*j*_ *−* 1,*T* + 1}, where asymptomatic infection and recovery before the study is labelled as (*A*_*j*_, *R*_*j*_) = (0, 0), asymptomatic infection before and recovery during/after the study is labelled as (*A*_*j*_, *R*_*j*_) = (0, *s*), (*s* 1,*…,T* + 1), and no asymptomatic infection before or during the study is labelled as (*A*_*j*_, *R*_*j*_) = (*T* + 1,*T* + 1). Posterior distributions for the asymptomatic infection and recovery time pairs are then estimated in the MCMC algorithm by proposing moves for these pairs and accepting/rejecting them based on the Metropolis-Hastings acceptance probability. The posterior distribution for which non-symptomatic individuals were asymptomatically infected during the study can be estimated from the proportion of time in the MCMC chain each individual spends in states other than (*T* + 1,*T* + 1). Since every non-symptomatic individual has an infection and recovery time pair, with a finite set of possible values, the dimension of the model remains fixed and reversible jump MCMC (RJMCMC) is not required. We note, however, that the algorithm described below for updating the asymptomatic infection and recovery time pairs is equivalent to a classical RJMCMC algorithm in which only individuals that are asymptomatically infected before or during the study have asymptomatic infection and recovery times and the number of asymptomatic infections varies as asymptomatic infection times are added or removed in the steps of the algorithm (31–34). As in the RJMCMC algorithm, the likelihood can be higher in the initial iterations of the chain than the value it ultimately converges to in our approach since the number of asymptomatic infections in the likelihood changes as the number of pairs corresponding to asymptomatic infection before or during the study changes, despite the fixed dimension of the parameter space.

In order to sample effectively from the posterior distribution of the asymptomatic infection and recovery times, we need to use a proposal distribution that allows us to efficiently navigate the very large space of possible asymptomatic infection and recovery time pairs. We therefore use the running estimate of the total infection pressure on each individual, averaged over the history of the MCMC chain, to propose new asymptomatic infection times for non-symptomatic individuals. The proposal distribution for individual *j* at the *k*th iteration of the MCMC chain thus consists of the probabilities of them being asymptomatically infected at each time point according to the mean infection pressure on them at time *t* from the previous samples in the chain,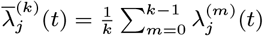:

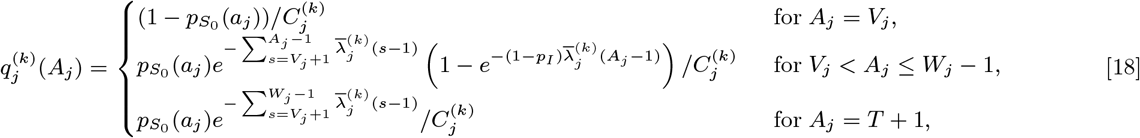

where 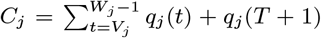 is a normalising constant to account for the fact that we know that *j* was not pre-symptomatically infected during the study. Although the infection pressure with a newly proposed asymptomatic infection and recovery time pair 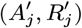 will be different from that with the current pair (*A*_*j*_, *R*_*j*_), and the probability of the reverse move 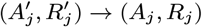 will therefore not be equal to the probability of the forward move 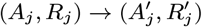, the difference in the infection pressure between consecutive iterations will be 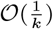. Hence, as the number of iterations becomes large, the adaptation will diminish (35) and 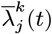 will tend towards a constant, such that the proposal probabilities for the forward and backward moves may be treated as equal.

#### Prior distributions

We use relatively weak Gamma distributions for the prior distributions for the transmission parameters *β, α, ϵ* and *δ*, which are non-negative, since there is little information available with which to construct informative priors (Table S3). The mean of the prior distribution for *α* is chosen as 100m based on our previous findings (1). A beta distribution, Beta(*a, b*), is chosen as a conjugate prior for the incubation period parameter *p*, since it is a probability (*p ∈* (0, 1]). The parameters of the beta distribution are chosen to match the mean of the prior distribution for the incubation period (NB(*r, p*), *p* Beta(*a, b*)) with the estimated mean incubation period (*∼*5 months) taken from previous analysis of diagnostic data from a subset of the study population (26). With this choice of prior the conditional posterior distribution of *p* is a beta distribution:

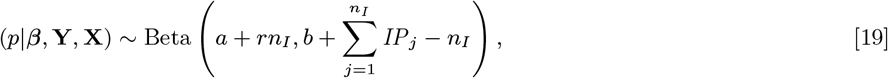

where ***β*** = (*β, α, ϵ, δ*), so *p* can be updated efficiently in the MCMC by drawing from this full conditional distribution rather than using a random walk Metropolis-Hastings update.

**Table S3.** Definitions and prior distributions of estimated parameters.

| Parameter (units) | Definition | Prior distribution |
| --- | --- | --- |
| $\beta$ (mnth<sup>-1</sup>) | Rate constant for spatial transmission from VL cases | Gamma(1, 1) |
| $\alpha$ (m) | Inverse distance decay rate of spatial kernel | Gamma(1, 100) |
| $\epsilon$ (mnth<sup>-1</sup>) | Background transmission rate | Gamma(1, 1) |
| $\delta$ | Additional within-household transmission rate | Gamma(1, 1) |
| $p$ | Incubation period distribution parameter | Beta(22, 28) |

#### Initial parameter values and missing data values

Informed guesses based on previous results (1) and preliminary runs of the MCMC are used for the starting values for the model parameters to reduce the burn-in time, ***θ***_0_ = (*β*_0_, *α*_0_, *ϵ*_0_, *δ*_0_, *p*_0_) = (3, 100, 0.001, 0.001, 3*/*7). The initialisation of the missing data, **X**, is more involved as it requires picking starting values for the asymptomatic “infection” and “recovery” times of all non-symptomatic individuals in the population (which determine their initial statuses and whether they were infected or not by the end of the study), along with the infection times of all VL cases, and proceeds as follows:

- For the VL case missing a treatment time, draw a treatment time using Eq. (5), i.e.

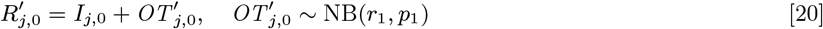
- For each VL case *j* missing a treatment time who may have had active VL at the start of the study (assumed to be only those who had onset after January 2001):

– if their onset time *I*_*j*,0_ is missing, draw it from *U* (*−*11, 0) (i.e. uniformly from January 2001–December 2001)
– draw their treatment time *R*_*j*,0_ as in Eq. (20)
- For each VL case *j* that suffered relapse:

– if their relapse time *I*_*Rj*,0_ is missing, draw it using Eq. (8)
– draw their relapse treatment time *R*_*Rj*,0_ using Eq. (5), i.e.

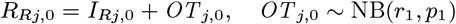
- For each VL case *j*, draw an infection time

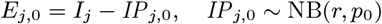
- For each non-symptomatic individual *j*:
  – draw their initial status (susceptible, currently asymptomatically infected, or previously asymptomatically infected and recovered) according to Eq. (13)–(15)
  – if they are initially recovered from previous asymptomatic infection, set (*A*_*j*,0_, *R*_*j*,0_) = (0, 0)
  – if they are initially actively asymptomatically infected, draw their recovery time *R*_*j*,0_ from 1,*…, W*_*j*_ *−* 1,*T* + 1 with probability

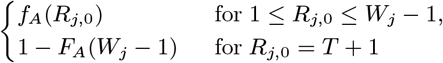
  – for the subset that are initially susceptible:
    ∗ calculate the infection pressure on them (Eq. (9)) from VL cases and PKDL cases with prior VL
    ∗ use this to calculate the probability of asymptomatic infection at each time point as in Eq. (18), i.e. with 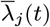 replaced by the infection pressure from VL and PKDL cases
    ∗ draw 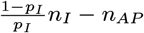 individuals, where *n*_*AP*_ is the number of asymptomatic individuals who develop PKDL, to be asymptomatically infected according to their cumulative probability of asymptomatic infection before the end of the study, 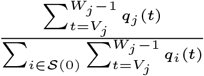
    ∗ for each asymptomatic individual draw an infection time *A*_*j*,0_ *∈ {V*_*j*_ + 1,*…, W*_*j*_ *−* 1*}* with probability

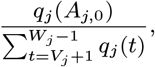

and a recovery time *R*_*j*,0_ *∈ {A*_*j*_ + 1,*…, W*_*j*_ *–* 1,*T* + 1*}* with probability

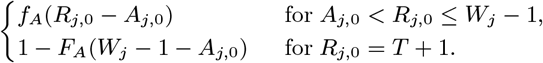
    ∗ for all individuals not infected before the end of the study set (*A*_*j*,0_, *R*_*j*,0_) = (*T* + 1,*T* + 1)
- For each PKDL case *j* without prior VL, set a dormant infection start time, *D*_*j*,0_, by drawing a dormant infection duration and subtracting it from their PKDL onset time,

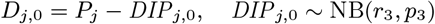

and then draw an asymptomatic infection time, *A*_*j*,0_, conditional on *A*_*j*,0_ *> V*_*j*_,

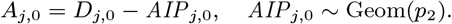

The MCMC algorithm was run from a range of initial parameter values and asymptomatic infection and recovery time configurations (with different numbers of individuals asymptomatically infected before the study and during the study etc.) to test convergence, and in all cases converged to the same posterior distributions.

#### MCMC algorithm

With the initial parameter values, missing data and priors chosen as described, the MCMC algorithm proceeds by repeating the following steps. Note that throughout the following we suppress notation of conditional dependencies in the likelihood terms where they are obvious to maintain legibility. The algorithm also accounts for the fact that some individuals were born or migrated or died during the study when updating the unknown pre-symptomatic infection times and asymptomatic infection and recovery times (using the birth/migration/death times as bounds on the proposed unobserved times), but we omit these details from the following description for simplicity.

##### 1. Update transmission parameters

Update the transmission parameters ***β*** = (*β, α, ϵ, δ*) conditional on the current incubation period distribution parameter *p* and missing data values **X** and observed data **Y**, using an adaptive random walk Metropolis-Hastings step:

a. Propose new values ***β*** as described in *Accelerated adaptive random walk Metropolis algorithm* below.
b. Accept ***β*** with probability

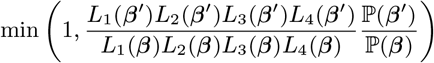

##### 2. Update incubation period distribution parameter

Update *p* by drawing from its conditional posterior distribution ℙ (*p*|***β*, Y, X**) (Eq. (19)).

##### 3. Move pre-symptomatic infection times

Update one-at-a-time the pre-symptomatic infection times of a random 20% of VL cases for whom both the onset and treatment times were observed, and the infection times of all cases missing their onset and/or treatment time:

a. For each case *j*, propose a new infection time conditional on the current values of the other infection times and the model parameters, 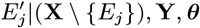, **Y, *θ***, using an independence sampler, i.e. propose a new incubation period 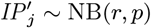 and subtract this from the onset time

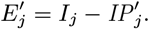
b. Accept the infection time move with probability

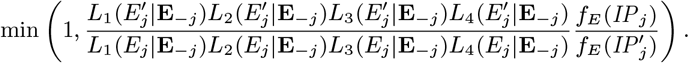

where 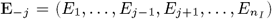 is the set of pre-symptomatic infection times with *j*’s infection time removed.

##### 4. Update asymptomatic infection and recovery times

Update one-at-a-time the asymptomatic infection and recovery times of 20% of non-symptomatic individuals as follows:

a. Choose a non-symptomatic individual *j* with probability proportional to their cumulative probability of being asymptomatically infected before the end of the study 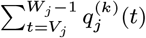.
b. Draw a new asymptomatic infection time 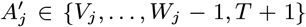 from the asymptomatic infection time proposal distribution 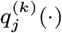 (Eq. (18)).
c. i. If *A*_*j*_ = 0: If 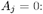, propose a new 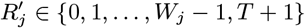 with probability

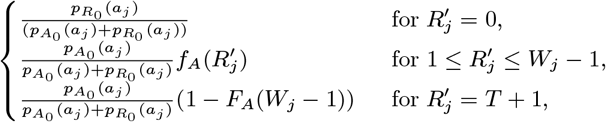

Accept 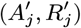 with probability

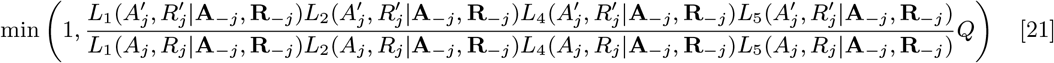

where

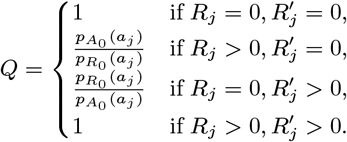

and **A**_*−j*_ = **A** *\ {A*_*j*_*}* and **R**_*−j*_ = **R** *\ {R*_*j*_*}*. B. If 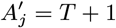, set 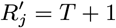 and accept 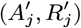 with probability in Eq. (21), but where *Q* is now

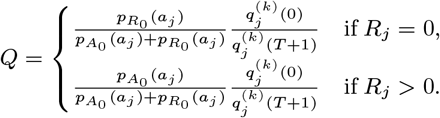

C. If 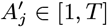, propose 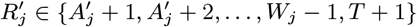 with probability

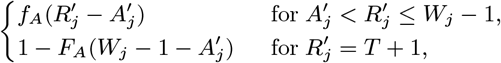

and accept 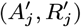 with probability in Eq. (21) but with

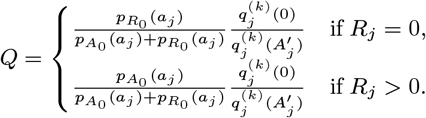 ii. If *A*_*j*_ = *T* + 1: A. If 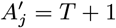, then 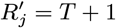, so accept immediately as the likelihood does not change. B. If 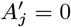, follow Step 4(c)iA, except with

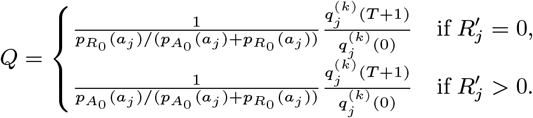 C. If 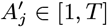, follow Step 4(c)iC, except with

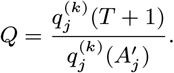 iii. If *A*_*j*_ *∈* [1,*T*]: A. If 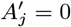, follow Step 4(c)iA, but with *Q* replaced by

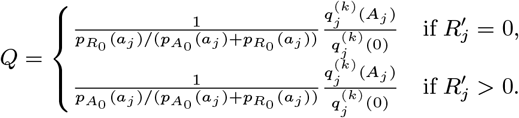

B. If 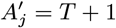, follow Step 4(c)iB except with

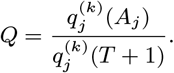

C. If 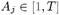, follow Step 4(c)iC but with

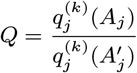

##### 5. Update asymptomatic infection times and dormant infection times of PKDL cases without prior VL

Update the asymptomatic infection and dormant infection times for each PKDL case *j* without prior VL, conditional on the asymptomatic infection time being after the start of the study or *j*’s birth (whichever is later):

a. Propose a new time, 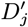, for recovery to dormant infection from asymptomatic infection using an independence sampler

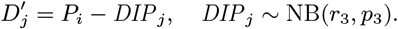
b. Propose a new asymptomatic infection time, 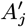

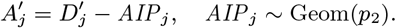
c. If 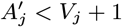, reject immediately. Otherwise accept 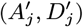 with probability

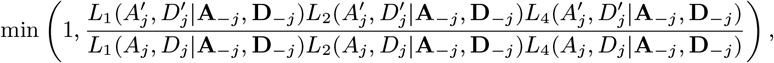

where **D**_*−j*_ = **D** *\ {D*_*j*_*}*.

##### 6. Update missing treatment times of VL cases during the study

Update the treatment time of the VL case whose treatment time is missing but whose onset time is known, conditional on the treatment time being before their PKDL onset:

a. Propose a new treatment time as

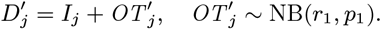
b. If 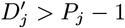, reject immediately. Otherwise accept 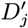 with probability min

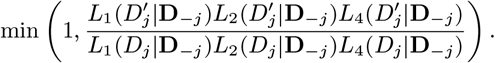

##### 7. Update whole onset-to-treatment period of potentially initially active VL cases

Update the onset and treatment times of all cases who potentially had active VL at the start of the study (*t* = 1) who are missing both onset and treatment times, one case at a time. For each case *j*:

a. Propose new onset and treatment times, 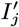 and 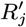

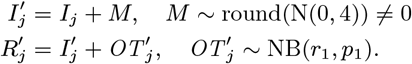
b. If 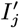 is not in *j*’s onset year or 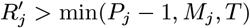, reject immediately. Otherwise, accept the new times with probability

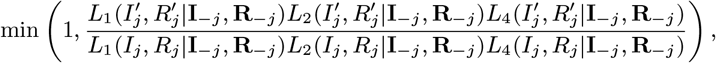

where **I**_*−j*_ = **I** *\ {I*_*j*_*}*.

##### 8. Update missing treatment times of potentially initially active VL cases

Update the treatment times of cases who potentially had active VL at the start of the study whose treatment times were not recorded but whose onset times are known, one by one. For each case *j*:

a. Propose a new treatment time, 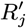

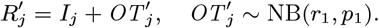
b. If 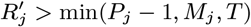, reject immediately. Otherwise, accept 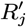 with probability

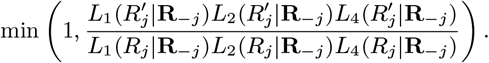

##### 9. Update whole relapse period of cases missing both relapse and relapse treatment times

Update the relapse and relapse treatment times of all VL cases who suffered relapse during the study who are missing both times, one case at a time. For each case *j*:

a. Propose new relapse and relapse treatment times, 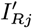 and 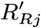

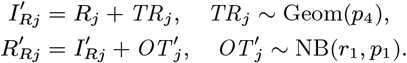
b. If 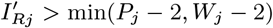 or 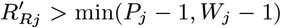, reject immediately. Otherwise, accept 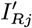 and 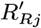 with probability

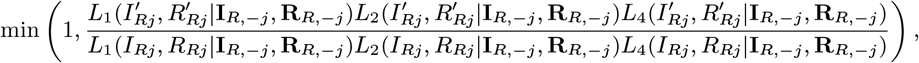

where ***I*** _*R, − j*_ = ***I***_*R*_ \ {***I*** _*Rj*_} and ***R*** _*R, − j*_ = ***R*** _*R*_ \ {*R* _*Rj*_}

##### 10. Update missing relapse treatment times of relapse VL cases

Update the missing relapse treatment times of VL cases with known relapse times:

a. Propose a new relapse treatment time, 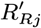

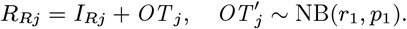
b. If 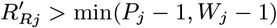, reject immediately. Otherwise, accept 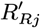 with probability

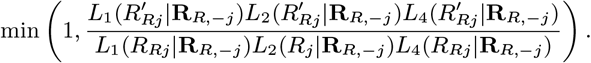

Terms in the full likelihood (Eq. (16)) for the asymptomatic infection duration, dormant infection duration, VL symptom duration of cases with missing onset and/or treatment time, and relapse duration do not appear in the acceptance probabilities for the updates for asymptomatic infection and recovery times, dormant infection times, missing onset and treatment times, and missing relapse and relapse treatment times (steps 4–10), as they cancel with the proposal probabilities for the newly proposed times, and are unaffected by the other steps in the algorithm.

Step 4 in the above algorithm may appear complicated, but essentially consists of proposing a new asymptomatic infection time and recovery time pair 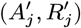 from anywhere in an approximately triangular grid of possibilities in discrete (*A, R*)-space (since *R*_*j*_ *≥ A*_*j*_ with equality only when *A*_*j*_ = 0 or *A*_*j*_ = *T* + 1), recalculating the parts of the likelihood that change with 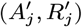 and accepting/rejecting according to the product of the likelihood ratio and the proposal ratio for the move 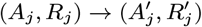.

We update 20% of infection times of VL cases and 20% of infection and recovery times of asymptomatic individuals in each iteration to strike a balance between efficient mixing of the MCMC chain and computational speed. The algorithm is run for *N* = 10^5^ iterations, including a burn-in of 20, 000 iterations which is discarded.

#### Accelerated adaptive random walk Metropolis algorithm

We block update the transmission parameters in step 1 of the algorithm above using a combination of the ‘Accelerated Shaping’ and ‘Accelerated Scaling’ algorithms described in (36). The Accelerated Shaping algorithm is a generalised version of the adaptive random walk Metropolis algorithm of Haario et al. (37), which uses the running estimate of the covariance matrix of the posterior distribution of the parameters from the MCMC chain in the multivariate normal proposal distribution to automatically tune the proposal covariance matrix to make efficient proposals. The Haario algorithm uses a fixed initial guess for the covariance matrix for a certain number of iterations at the start of the chain and then the estimate of the covariance matrix from the whole history of the chain after that, including the initial values and the burn-in, which can lead to slow convergence of the chain due to the sensitivity of the covariance estimate to outliers. The Accelerated Shaping algorithm instead begins adapting straight away and removes early iterations of the chain from the running estimate of the covariance matrix at a rate slower than it includes new iterations. This helps to ensure that the chain converges to the posterior mode quickly even if the initial guesses for the parameter values and covariance matrix are relatively poor.

If ***β***_*k*_ is the vector of the values of the transmission parameters at the *k*th iteration, then the proposal for ***β*** at the (*k* + 1)th iteration is given by

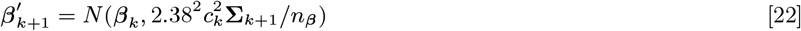

where **Σ**_*k*+1_ and *n*_***β***_ are the running estimate of the covariance matrix and dimension of the posterior density for ***β***, and *c*_*k*_ is a scaling constant that is tuned to achieve efficient mixing (see below). To discard early iterations at a slower rate than new ones are included, a non-decreasing sequence of integers is used, 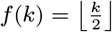, such that *f* (0) = 0 and *f* (*k* + 1) = *f* (*k*) or *f* (*k* + 1) = *f* (*k*) + 1 for all *k*, and **Σ**_*k*+1_ for *k >* 0 is defined as

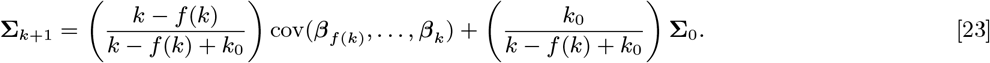

where

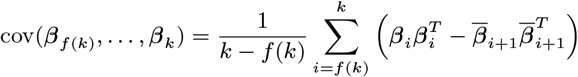

is the empirical covariance of the last *k − f* (*k*) + 1 samples of ***β*** from the chain, with

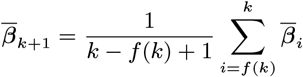

the mean of the last *k − f* (*k*) + 1 samples; **Σ**_0_ is the initial guess for the covariance matrix, and *k*_0_ determines the rate at which the influence of **Σ**_0_ on **Σ**_*k*+1_ decreases (the weight of **Σ**_0_ halves after the first 2*k*_0_ iterations). We use *k*_0_ = 1000 here.

If *f* (*k*) = *f* (*k −* 1) (i.e. if *k* is odd with *f* (*k*) chosen as above), an additional observation is added to the estimate of the covariance matrix

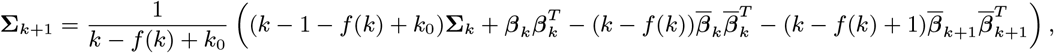

where

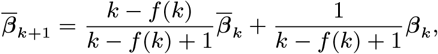

and if *f* (*k*) = *f* (*k −* 1) + 1 (i.e. if *k* is even), the new observation replaces the oldest

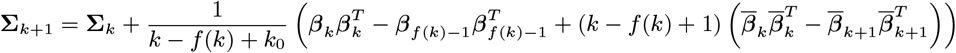

where

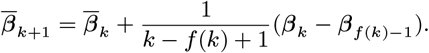

It has been shown that *N* (***β***_*k*_, 2.38^2^**Σ***/n*_***β***_), where **Σ** is the covariance matrix of the posterior distribution, is the optimal proposal distribution for rapid convergence and efficient mixing of the MCMC chain for symmetric product-form posterior distributions as *n*_***β***_ *→∞*, and leads to an acceptance rate of 23.4% (38, 39). This corresponds to a scaling of *c*_*k*_ = 1 in Eq. (23). However, we are in a context with a large amount of missing data, which is strongly correlated with some of the transmission parameters (see *Parameter estimates* below), so the posterior distribution is not symmetric, and this scaling is not optimal. We therefore follow (36) and scale *c*_*k*_ adaptively as the algorithm progresses to target an acceptance rate of approximately 23.4% for updates to ***β***. We do this by rescaling *c*_*k*_ by a factor of *x*_*k*_ *>* 1 every time an acceptance occurs and by a factor of 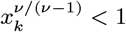 every time a rejection occurs such that the acceptance rate *ν* approaches 23.4% in the long run,

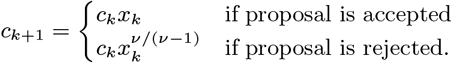

In order to satisfy the ‘Diminishing Adaptation’ condition (40), which is necessary to ensure the Markov chain is ergodic and converges to the correct posterior distribution, it is required that *c*_*k*_ tends to a constant as *k →∞*. So that the adaptation diminishes as *k* increases, we use the sequence

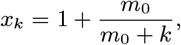

where *m*_0_ is the number of iterations over which the scaling factor *x*_*k*_ decreases from 2 to 1.5. Here, we use *m*_0_ = 100.

### Model comparison

We compare the goodness of fit of models with different asymptomatic and pre-symptomatic relative infectiousness (between 0% and 2% of that of VL cases), with and without additional within-household transmission, to test different assumptions about how infectious asymptomatic and pre-symptomatic individuals are, using DIC (41). DIC measures the trade-off between model fit and complexity and lower values indicate better fit. Since some variables were not observed, we use a version of DIC appropriate for missing data from (42), which is based on the complete data likelihood *L*(***θ***; **Z**) = ℙ (**Y, X**|***θ***). This is equivalent to the standard version of DIC for fully observed data except that it is averaged over the missing data:

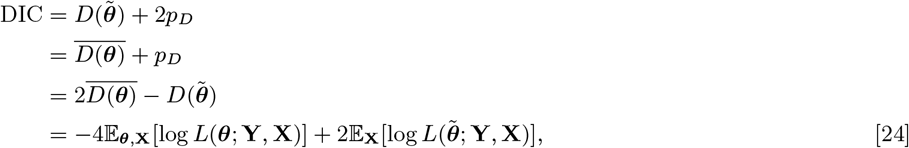

where *D*(***θ***) is the deviance (the measure of model fit), given (up to an additive constant dependent only on the data) by

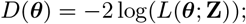

*p*_*D*_ is the effective number of parameters in the model (the measure of model complexity), defined by

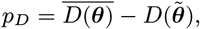

and 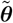 is an appropriate summary statistic for ***θ*** from its posterior distribution (here we use the posterior mode).

The first term in Eq. (24) can be straightforwardly estimated as the mean of the values of the log-likelihood over the MCMC samples:

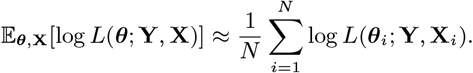

The second term in Eq. (24) is calculated by storing the values of the missing data while running the MCMC, re-running the full log-likelihood computation for each iteration using the stored values and the posterior mode, and averaging these values

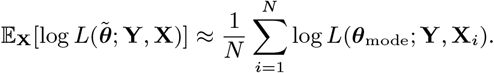

Since there can be issues with using DIC for model comparison (43), we also compared the posterior distributions of the deviances of the models to assess differences in quality of fit.

### Calculating the contribution of different infection states to transmission

We calculate the contribution of each infection state to the total infection pressure on all susceptible individuals *λ*(*t*), i.e. the total risk of new infections at time *t* (Fig. 2A in the main text), by summing the infection pressures from each state on susceptible individuals

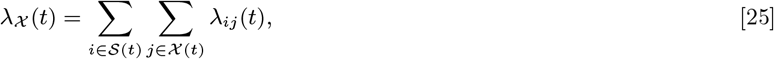

where *𝒳 ∈* {*𝒜,εℐ, 𝒫*} denotes the infection state. The relative contribution to the infection pressure on susceptible individuals (Fig. S17) is then given by

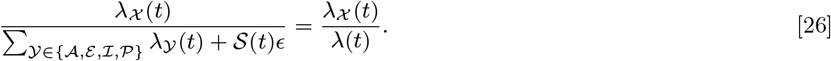

The relative contribution of state *𝒳* to the infection pressure on the *i*th VL case at their infection time, i.e. the probability that *i*’s infection source is *𝒳* (Fig. 2B in the main text), is:

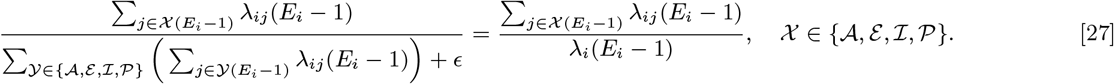

The probability that the *i*th VL case is infected from the background transmission is 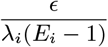.

### Reconstructing the epidemic

#### Reconstructing the transmission tree

We reconstruct the transmission tree following the ‘sequential approach’ described in (44). We draw *N* samples (***θ***_*k*_, **X**_*k*_) (*k* = 1,*…,N*) from the joint posterior distribution from the MCMC, calculate the probability that infectee *i* was infected by individual *j* conditional on their infection time *E*_*i*_

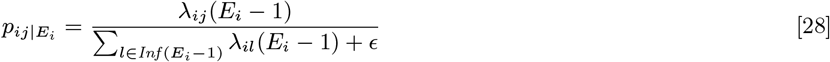

for all infectees and possible infectors for each sample, and draw an infector for each infectee from the posterior distribution of possible infectors *p*_*ij*|*E*_ (***θ***_*k*_, **X**_*k*_) from each sample. This yields a sample of *N* transmission trees drawn from their predictive distribution, and thus accounts for uncertainty in the infection source (given fixed values of the parameters and missing data), and uncertainty in the parameter values and missing data (over the posterior distribution). We use *N* = 1000 here.

#### Calculating transmission distances and times

The mean infector-to-VL-infectee distance and mean infector-onset-to-VL-infectee-infection time for each VL and PKDL infector (Figures 4A and 4B in the main text) are calculated from the sample of N transmission trees by averaging the distances and times from each infector to their VL infectees within each tree, and then averaging these quantities over all the trees in which that VL/PKDL case is an infector:

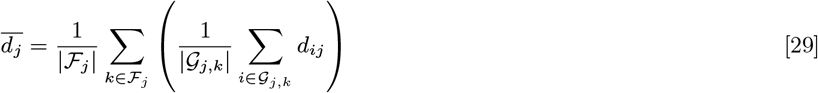

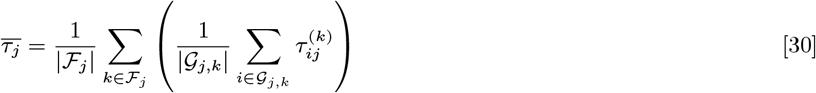

where 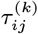 is the time between *j*’s onset and *i*’s infection in the *k*th tree, *ℱ*_*j*_ is the set of all trees in which *j* is an infector, *𝒢*_*j,k*_ is the set of all infectees of *j* in the *k*th tree, and |*·*| denotes set size.

#### Calculating reproduction numbers

The individual-level reproduction number for infectious individual *j* is obtained by summing the probabilities in Eq. (28) over all individuals *i* that *j* could have infected (45, 46):

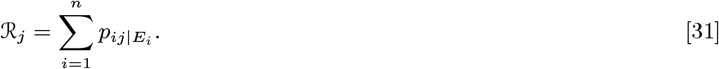

To account for uncertainty in infected individuals’ unobserved infection times these reproduction numbers are averaged over the joint posterior distribution of the transmission parameters and missing data:

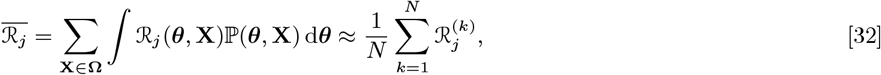

where **Ω** is the space of all possible values of the missing data 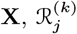 is individual *j*’s reproduction number from the *k*th iteration of the MCMC chain (with the burn-in removed), and *N* is a suitably large number of samples from the posterior distribution (we use *N* = 1000). Since we are interested in comparing the numbers of new infections generated by VL and PKDL cases, in the main text we separate the expected numbers of secondary infections coming from the VL episodes and PKDL episodes of individuals who had both VL and PKDL (Fig. 4C in the main text) (i.e. we effectively treat them as coming from separate individuals), and hence no longer refer to the numbers of secondary infections per VL case and per PKDL case as reproduction numbers.

The time-dependent overall effective reproduction number (Fig. S19C) can also be calculated from the transmission probabilities as the mean of the individual reproduction numbers for all VL cases and asymptomatic individuals with onset and infection, respectively, at time *t* (45):

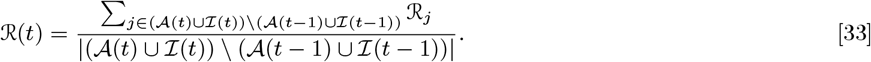

The absolute contribution of each infectious state to the effective reproduction number at time *t* is:

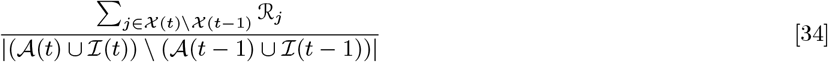

where *𝒳 ∈ {𝒜, ℐ}*, denotes the infectious state, and, as described above, in the main text we split the numbers of secondary infections (*ℛ*_*j*_) arising from VL and PKDL for cases that had both.

### Model simulations

To assess the fit of the model and simulate hypothetical interventions against PKDL, we create a stochastic simulation version of the individual-level spatiotemporal transmission model described above. We follow standard stochastic simulation methodology for discrete-time individual-level transmission models (47), converting infection event rates into probabilities in order to determine who gets infected in each month. We assume that an individual’s progression through different infection states following infection occurs independently of the rest of the epidemic (i.e. is either governed by internal biological processes or random external processes of detection), which enables the simulation of an individual’s full infection history from the point of infection.

So that we can simulate durations of PKDL infectiousness, we fit a negative binomial distribution NB(*r*_5_, *p*_5_) to the observed PKDL onset-to-treatment times and onset-to-resolution times for self-resolving PKDL cases in the data:

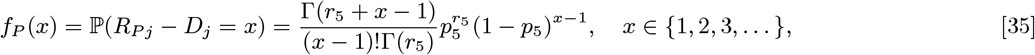

by maximum likelihood estimation, which yields *r*_5_ = 1.18, *p*_5_ = 0.066 (corresponding to a mean duration of PKDL infectiousness of 17.8 months).

We simulate epidemics for the study population starting from January 2003 (at which point all but one of the paras had had at least 1 VL case since January 2002) using the individual-level location and demographic information (months of birth, death and migration) from the observed data, and posterior samples of the parameter values and individuals’ infection statuses in December 2002 obtained from the MCMC algorithm.

Given these pieces of information, the simulation algorithm proceeds as follows:

1. Starting at *t* = 12, draw the times of the next events for all individuals that have already been infected, e.g. VL onset times for individuals already pre-symptomatically infected, using the size-biased negative binomial distributions corresponding to Eq. (4), Eq. (5), Eq. (7), and Eq. (35), and Eq. (6) for the asymptomatic infection durations. The size-biased negative binomial distribution accounts for the fact that individuals with left-censored infection times who are observed to be actively infected at a particular point in time are likely to have had long durations of infection if the infection duration is negative binomially distributed (48). The PMF of a size-biased negative binomial random variable *X*^*∗*^ corresponding to *X ∼* NB is:

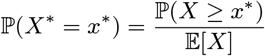
2. Simulate the times of the subsequent events for these individuals by drawing from Eq. (4)–Eq. (7) and Eq. (35). Assign each PKDL case an infectiousness by drawing from Cat(*h*_1_, *h*_2_, *h*_3_, *h*_*u*_, **p**), where Cat(·) is the categorical distribution and **p** = (101*/*190, 31*/*190, 6*/*190, 52*/*190) are the probabilities of the different lesion types according to their observed frequencies in the data.
3. For each susceptible individual *i* from the set of individuals who have been born/immigrated and have not yet died/emigrated, determine whether they become infected:
  a. Infection occurs with probability 1 *− e*^*−λi*(*t*)^, where *λ*_*i*_(*t*) is given by Eq. (9).
  b. Else they remain susceptible.
4. Let *t* = *t* + 1.
5. For each infection, decide if it is a pre-symptomatic infection or an asymptomatic infection:
  a. Pre-symptomatic infection occurs with probability *p*_*I*_. In which case, draw an onset time

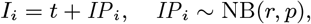

and decide whether the individual subsequently develops PKDL or not:
    i. PKDL occurs with probability *p*_*P*_. In which case draw times for progression to dormant infection, PKDL onset and recovery

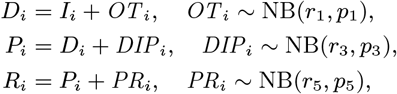

and assign a PKDL infectiousness to the individual by drawing from Cat(*{h*_1_, *h*_2_, *h*_3_, *h*_*u*_*}*, **p**).
    ii. Else the individual recovers upon treatment, so draw a treatment time

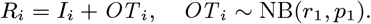
  b. Else it is an asymptomatic infection. In which case, decide whether the individual progresses to PKDL or not:
    i. Progression to PKDL occurs with probability *p*_*D*_. In which case, draw times for progression to dormant infection, PKDL onset and recovery

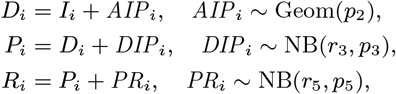

and assign a PKDL infectious by drawing from Cat(*{h*_1_, *h*_2_, *h*_3_, *h*_*u*_*}*, **p**).
    ii. Else the individual recovers without developing PKDL, so draw a recovery time:

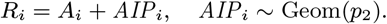
6. Repeat steps 3–5 until *t* = *T*.

We run 10,000 simulations of the model for each PKDL intervention scenario (normal interventions, complete PKDL prevention (*h*_1_ = *h*_2_ = *h*_3_ = *h*_*u*_ = 0) and halving the mean duration of infectiousness (*r*_5_ = 1.18, *p*_5_ = 0.13)), consisting of 100 simulations (to account for stochastic uncertainty) of each of 100 posterior samples for the model parameters and individuals’ infection statuses in December 2002 (to account for uncertainty in their values given the observed data).

### Computer code

Code for running the MCMC algorithm and processing the MCMC output was written in MATLAB R2017b (49) and the simulation code was written in Julia 1.0.5 (50, 51). All code is freely available from https://github.com/LloydChapman/ VLSpatiotemporalModelling.

### MCMC output

#### Model selection

The posterior deviance distributions and DIC values for the different models tested are presented in Figure S5 and Table S4 respectively. Based on the deviance distributions and DIC values, the best-fitting model is the model with additional within-household transmission and the highest level of relative pre-symptomatic and asymptomatic infectiousness (both 2% as infectious as VL). Hence, we focus on the output of this model in the main text and below.

**Fig. S5.**
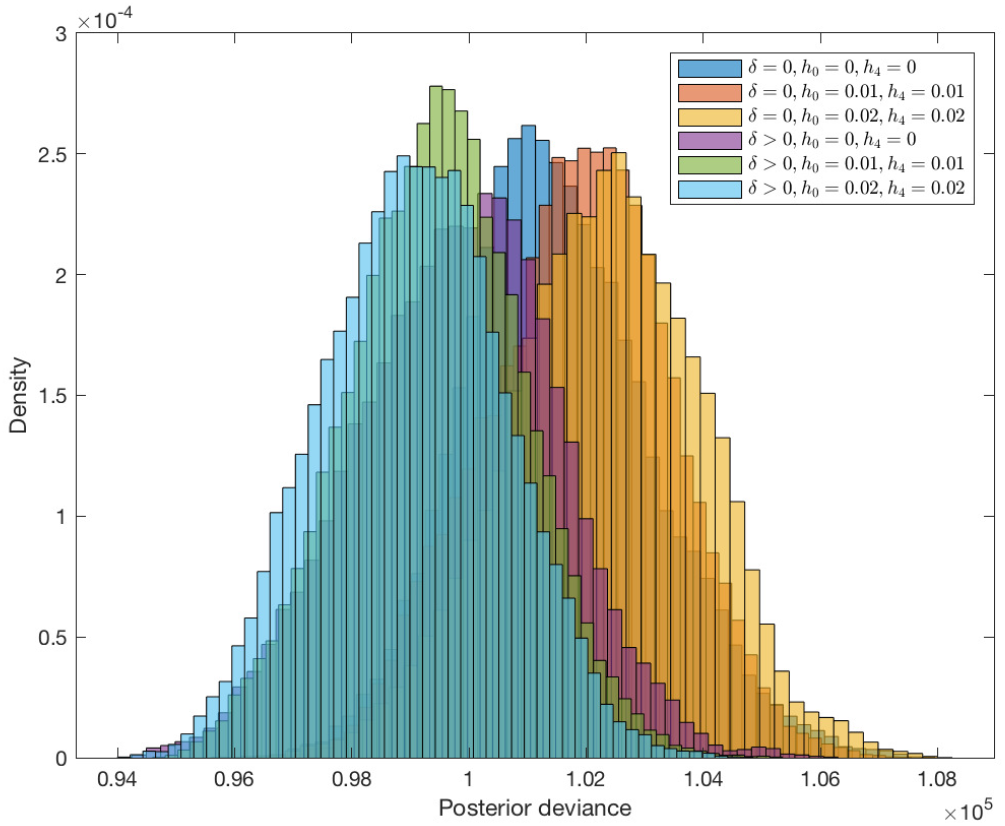
Posterior deviance distributions for different models tested

#### Parameter estimates

The modes and 95% highest posterior density intervals (HPDIs) of the transmission parameters (*β, α, ϵ* and *δ*) and incubation period distribution parameter *p* for the different models are shown in Table S4. The parameter estimates are very similar across the different models and vary in the way expected – the spatial transmission rate constant *β* and background transmission rate *∈* are lower for models with additional within-household transmission (*δ >* 0) and decrease with increasing relative asymptomatic infectiousness *h*_4_, and the mode for *α* is slightly larger for models with *δ >* 0 (since a flatter kernel shape compensates for the extra within-household transmission). The posterior distributions for the incubation period distribution parameter *p* correspond to a mean incubation period of 5.7–6.9 months (95% HPDIs (4.8,6.6)–(6.0,7.8) months).

**Fig. S6.**
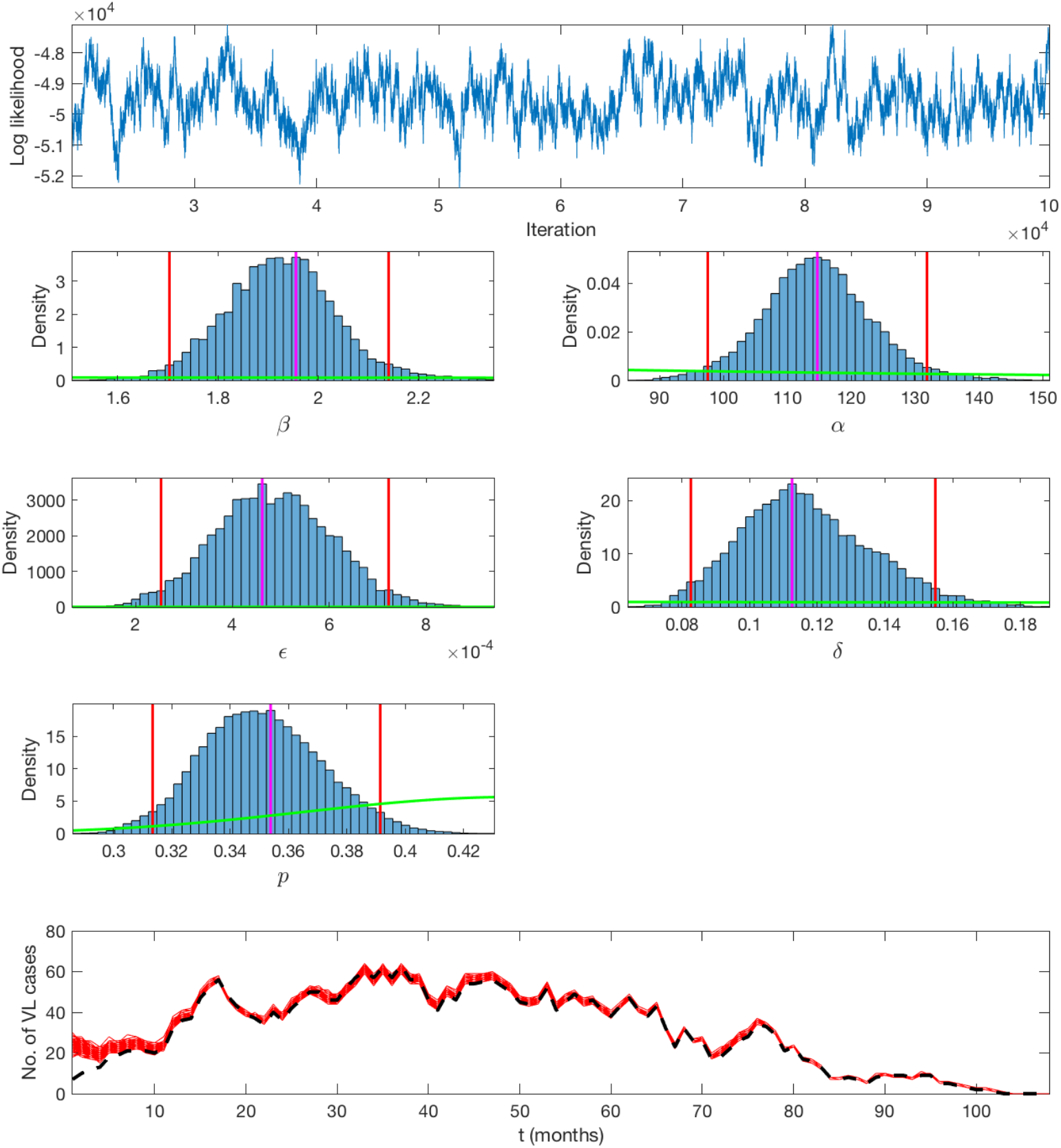
Output of the MCMC algorithm for the best-fitting model with additional within-household transmission and 1% asymptomatic infectiousness relative to VL cases. Top: Log-likelihood trace plot. 2nd-4th row: Posterior distributions for the spatial transmission rate constant *β* (mnth^*−*1^), risk decay distance *α* (m), background transmission rate (mnth^*−*1^), additional within-household transmission rate *δ* (mnth^*−*1^), and incubation period distribution parameter *p*, with prior distributions (green lines), posterior modes (magenta lines) and 95% highest posterior density intervals (red lines). Bottom: Number of active VL cases over time in 100 iterations of the MCMC algorithm (red lines), with inferred missing onset and recovery times, and number of active cases excluding individuals with missing onset and/or recovery times (black dashed line).

The log-likelihood trace and posterior distributions for the parameters for the best-fitting model are shown in Figure S6. The parameters are clearly well defined by the data, as the posterior distributions differ significantly from the weak prior distributions.

The corresponding autocorrelation plots are shown in Figure S7. The high degree of autocorrelation evident for all the parameters is due to strong correlation between the transmission parameters and the missing data, in particular between the spatial transmission rate constant *β* and the asymptomatic infection times. Figure S8 shows that *β* is strongly negatively correlated with the mean asymptomatic infection time 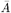 This is expected since a higher overall transmission rate leads to individuals being infected earlier. With a continuous-time model we could tackle this correlation using a ‘non-centred’ MCMC algorithm, i.e. by re-parameterising the model to reduce the *a priori* dependence between *β* and the asymptomatic infection times (30, 33), but with a discrete-time model the re-parameterisation is much more difficult, since the asymptomatic infection times do not vary continuously with *β*. We therefore ran the MCMC algorithm for a large number of iterations (*N* = 10^5^) to obtain a sufficient number of independent samples.

Correlations between the model parameters were explored by plotting the pairwise marginal posterior densities (Figure S9) and calculating the correlation coefficient matrix of the MCMC samples, 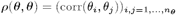, where corr 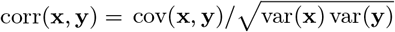, and var(**x**) is the sample variance of **x** and *n*_***θ***_ is the dimension of ***θ***. Figure S9 and the correlation coefficient matrix,

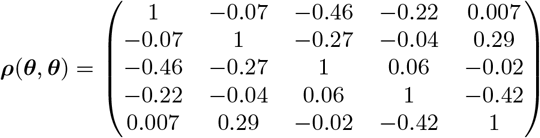

show that there is some negative correlation between *β* and *ϵ, α* and *ϵ*, and *δ* and *p*. These correlations are not surprising: the more transmission that is explained by proximity to infectious individuals (the higher *β*), the less needs to be explained by the background transmission (the lower *ϵ*); the flatter the spatial kernel (the larger *α*), the fewer infections need to be explained by the background transmission; and the more infections are accounted for by transmission within the same household (the higher *δ*), the longer the incubation period (the lower *p*) needs to be (due to long times between onsets of cases in the same household).

**Table S4.** Parameter estimates (modes and 95% highest posterior density intervals) for models with different relative pre-symptomatic and asymptomatic infectiousness, *h*_0_ and *h*_4_, and without and with additional within-household transmission, *δ* = 0 vs *δ >* 0

| Parameter | No additional within-household transmission, $\delta = 0$ | | | Additional within-household transmission, $\delta > 0$ | | |
| --- | --- | --- | --- | --- | --- | --- |
| | $h_0, h_4 = 0$ | $h_0, h_4 = 0.01$ | $h_0, h_4 = 0.02$ | $h_0, h_4 = 0$ | $h_0, h_4 = 0.01$ | $h_0, h_4 = 0.02$ |
| $\beta$ (mnth<sup>-1</sup>) | 2.60 (2.31,2.87) | 2.52 (2.24,2.74) | 2.38 (2.14,2.64) | 2.24 (1.97,2.48) | 2.00 (1.81,2.28) | 1.96 (1.70,2.14) |
| $\alpha$ (m) | 105 (89,120) | 103 (88,119) | 97 (86,113) | 121 (103,139) | 117 (102,137) | 115 (98,132) |
| $\epsilon$ (10<sup>-4</sup> mnth<sup>-1</sup>) | 5.9 (3.7,8.8) | 5.2 (3.2,8.6) | 5.6 (3.3,8.2) | 5.1 (3.4,8.6) | 5.2 (3.2,8.1) | 4.6 (2.5,7.2) |
| $\delta$ (mnth<sup>-1</sup>) | 0 | 0 | 0 | 0.10 (0.06,0.13) | 0.10 (0.08,0.15) | 0.11 (0.08,0.15) |
| $p$ | 0.41 (0.37,0.46) | 0.41 (0.36,0.46) | 0.40 (0.36,0.46) | 0.37 (0.34,0.41) | 0.36 (0.32, 0.40) | 0.35 (0.31,0.39) |
| ΔDIC | 2512 | 2974 | 3264 | 948 | 470 | - |

**Fig. S7.**
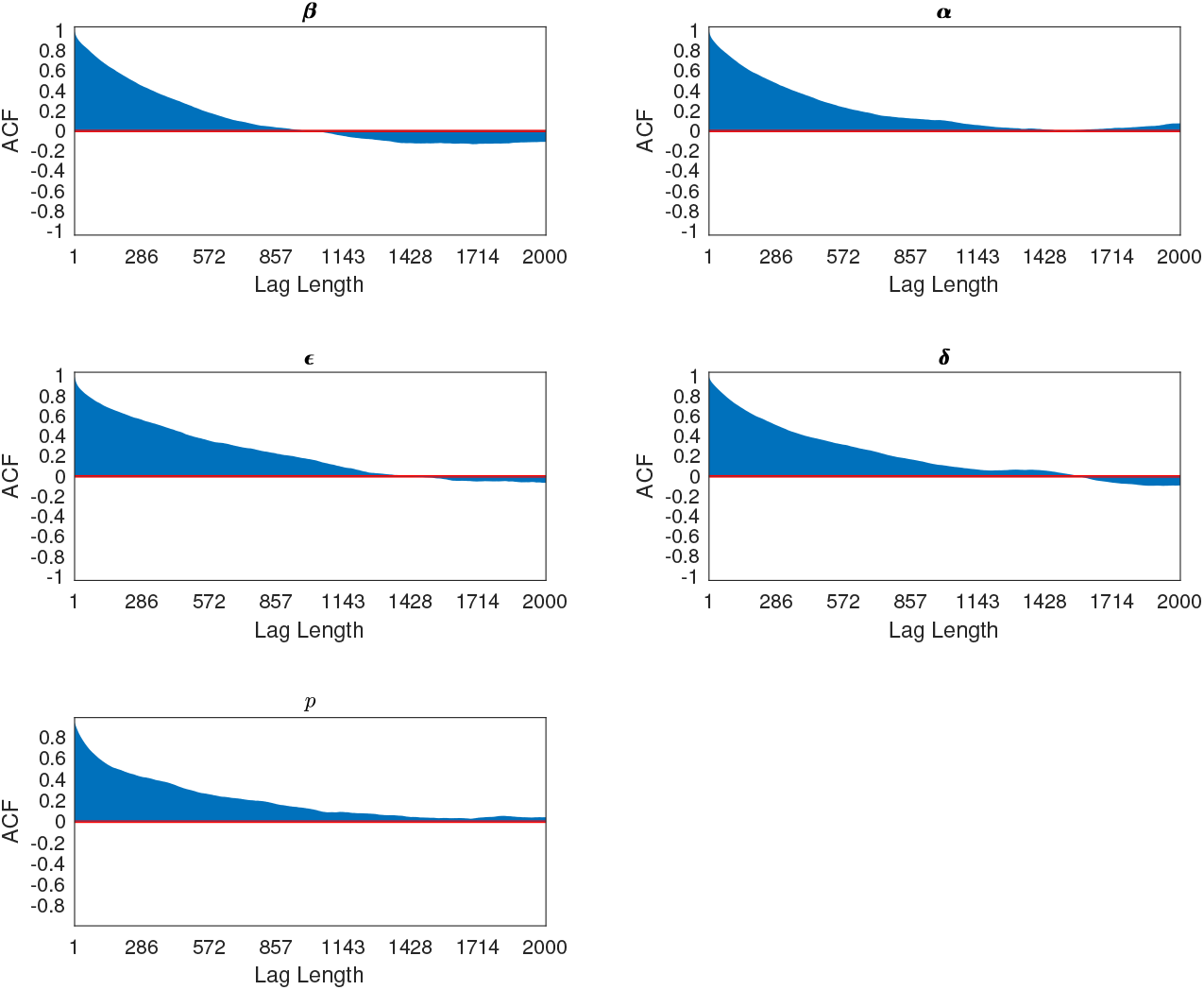
Autocorrelation plots for the model parameters ***θ*** = (*β, α, ϵ, δ, p*) for the best-fitting model, showing the correlation between samples in the MCMC chain at different lags.

The acceptance rate for the transmission parameter updates (Step 1 in the MCMC algorithm) was 23.3–23.4% for all models, as expected from tuning the proposal variance (see *Accelerated adaptive random walk Metropolis algorithm*). The acceptance rates for the updates for the pre-symptomatic infection times; missing VL onset and treatment times; and unobserved asymptomatic infection and recovery times were between 57% and 97% for every model, indicating that the algorithm efficiently explores the space of unobserved data.

**Fig. S8.**
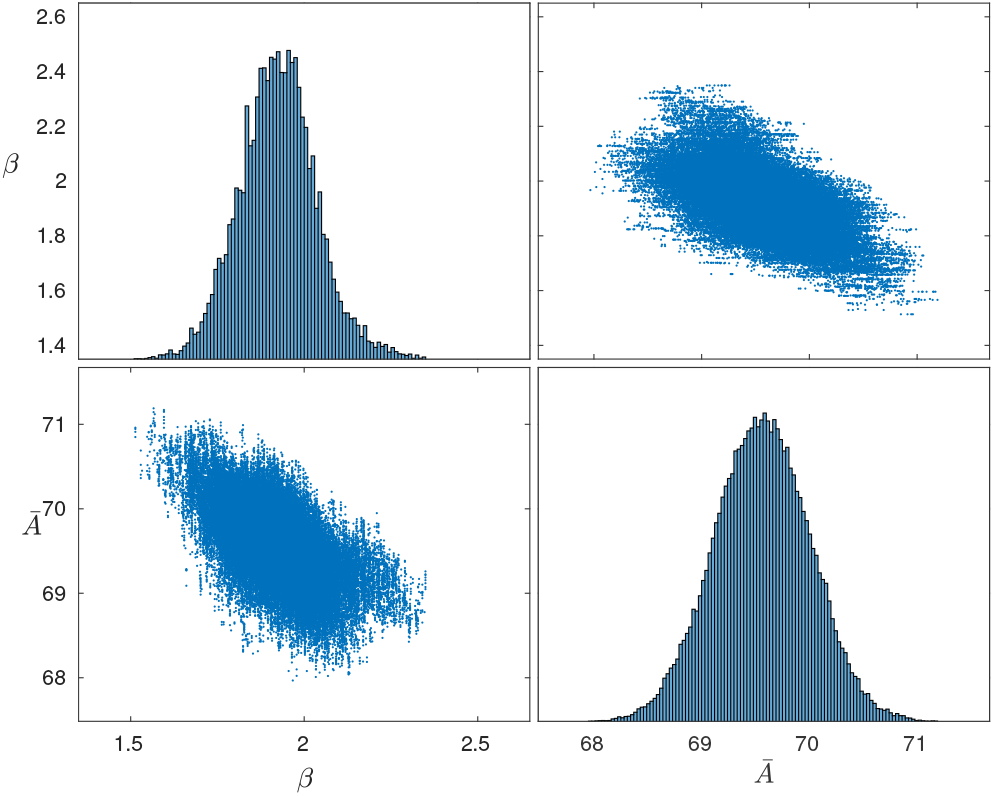
Correlation between the spatial transmission rate constant *β* and the mean asymptomatic infection time 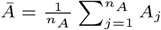, where *n*_*A*_ = |*{j* : *A*_*j*_ *< T}*| is the number of individuals asymptomatically infected before the end of the study.

**Fig. S9.**
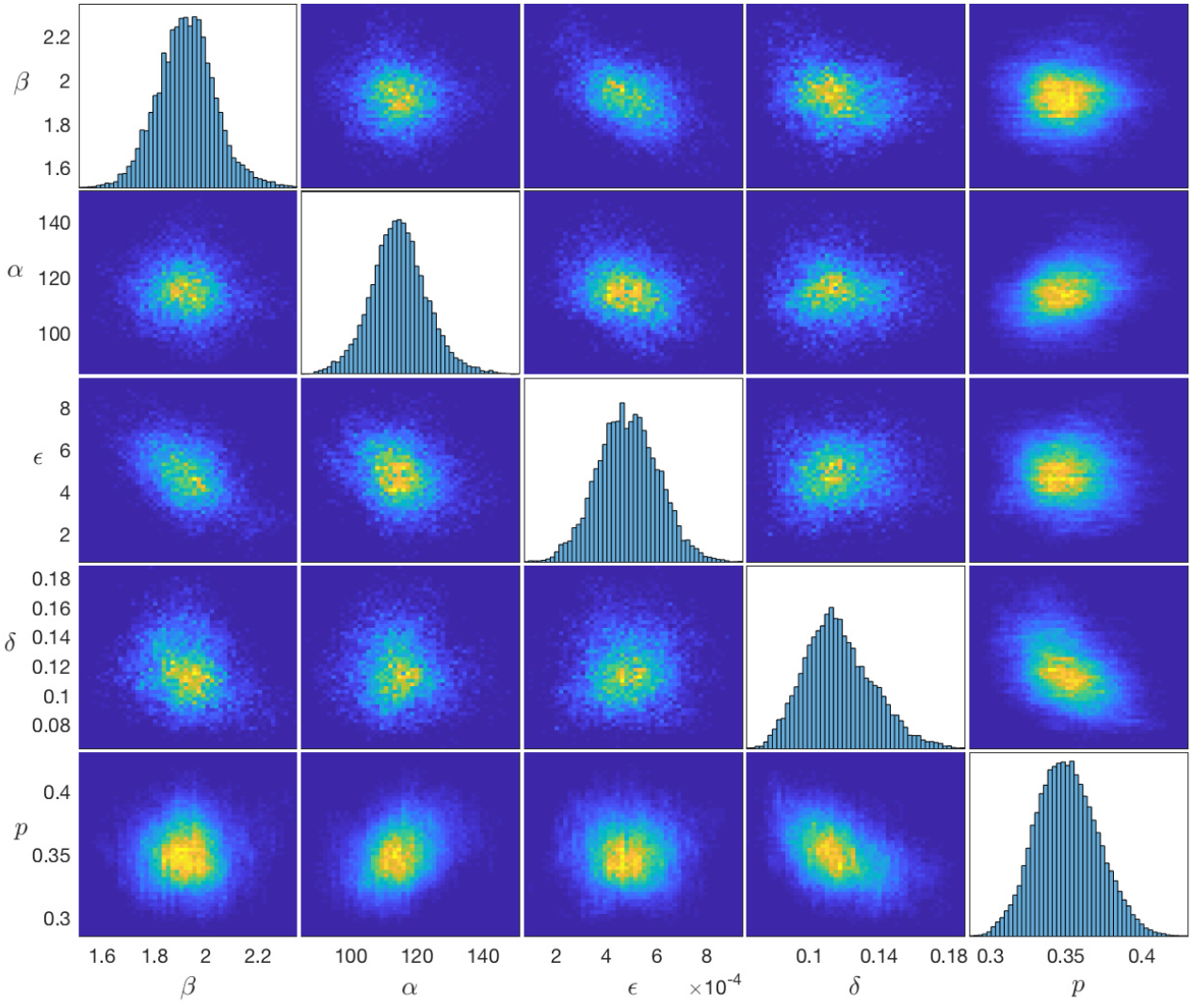
Posterior distributions and pairwise correlations of the transmission parameters for the best-fitting model. Main diagonal: Marginal posterior distributions of the transmission parameters. Off diagonals: Heatmap plots of pairwise marginal posterior densities showing the correlations between parameters. Lighter yellower colours indicate areas of higher posterior density.

#### Unobserved pre-symptomatic infection times, and asymptomatic infection and recovery times

Here we present plots of various quantities derived from the inferred pre-symptomatic infection times and asymptomatic infection and recovery times to demonstrate that the data augmentation algorithm works as expected. Figure S10 shows the incidence curve of VL and PKDL cases for the whole study area and the inferred incidence curve of asymptomatic infections (averaged over the MCMC chain). The number of asymptomatic infections increases and decreases with the number of VL cases as expected given the assumption that the incidence ratio of asymptomatic to symptomatic infection is fixed.

The posterior probabilities that individuals were asymptomatically infected during the study (shown in Figure S11, with individuals arranged in order of para and household number) show clustering of asymptomatic infection in space and that this is associated with proximity to VL cases (the probabilities of asymptomatic infection are higher where the VL case density is higher). This is as expected given the structure of the model (the decrease in the risk of infection with distance from an infectious individual encoded in the spatial kernel) and the estimates of the transmission parameters.

**Fig. S10.**
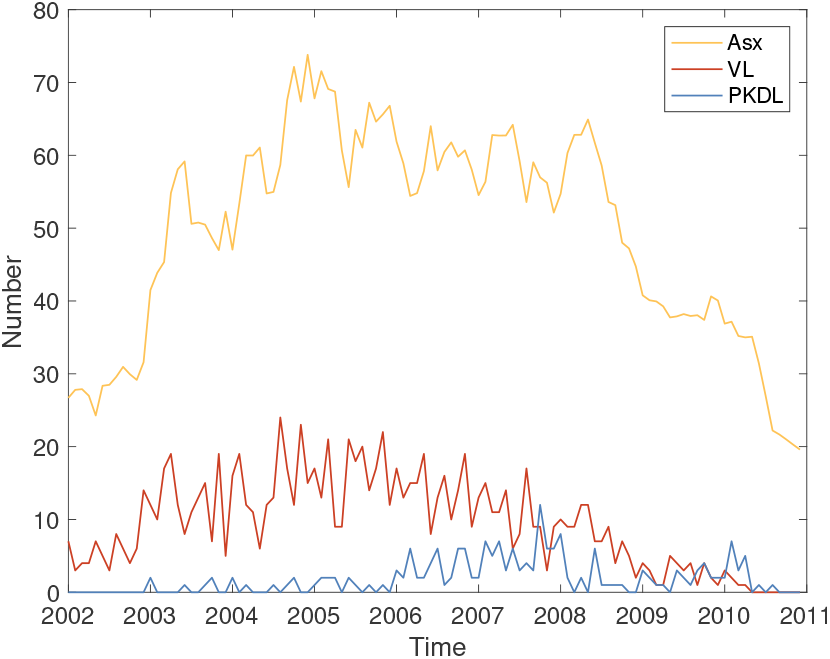
Numbers of new VL and PKDL cases over time in the study area, with inferred numbers of new asymptomatic infections.

**Fig. S11.**
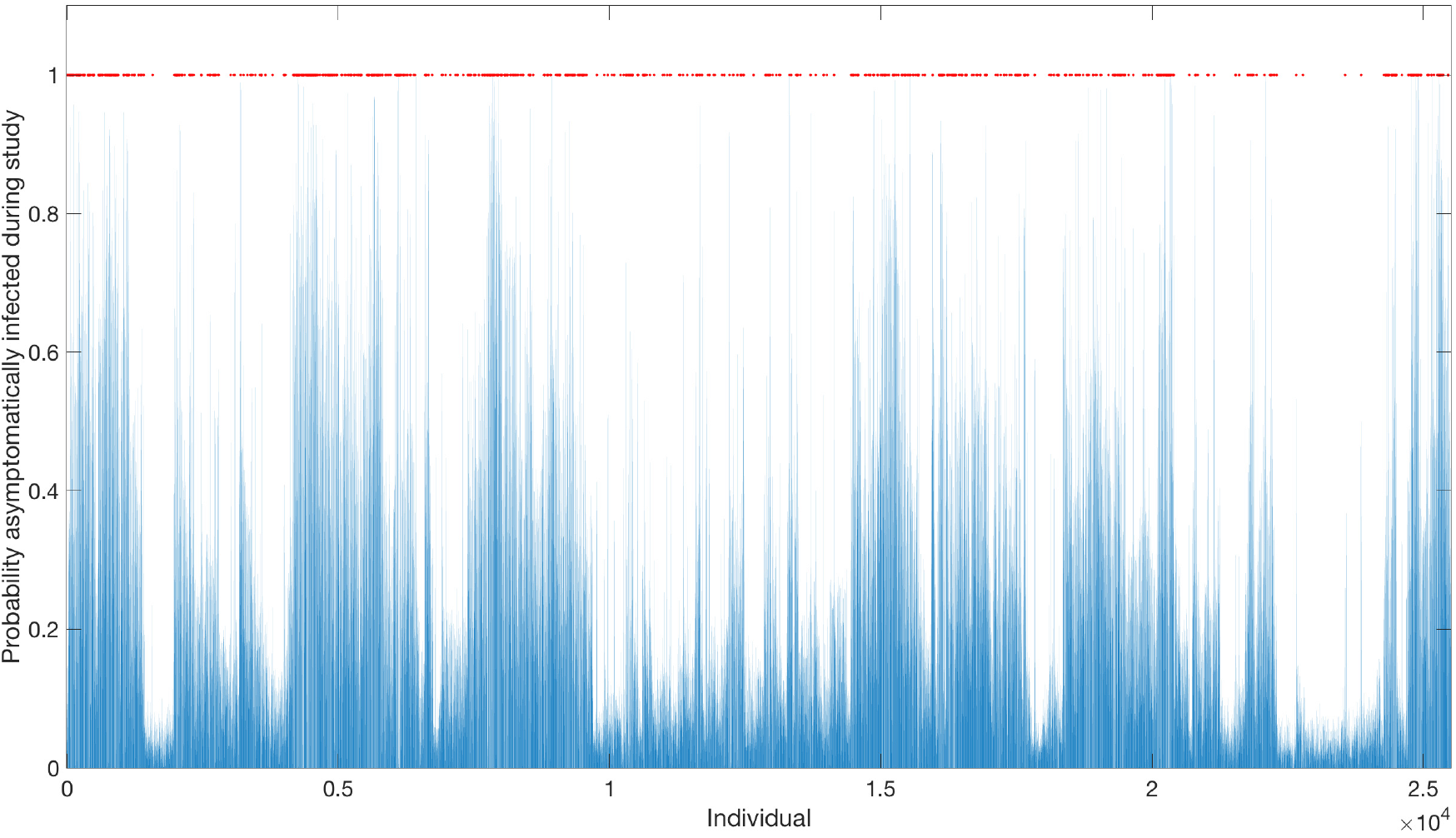
Posterior probabilities individuals were asymptomatically infected during the study (blue lines). Individuals who had VL onset during the study are marked by red dots.

The examples shown in Figure S12 demonstrate that non-symptomatic individuals’ asymptomatic “infection” time posterior distributions and asymptomatic infection time proposal distributions (probabilities of asymptomatic infection according to the empirical average of the infection pressure on them over the history of the chain) converged with each other. This is expected since the model parameters converge to the high posterior density part of the parameter space, such that the running average of the infection pressure on each individual at each time point approaches a constant and the asymptomatic infection times are effectively sampled from their posterior distributions in the long run. The plots also show that the data augmentation algorithm accounts for individuals’ birth, migration and death times when updating their asymptomatic infection time.

Figure S13 shows the posterior distributions of the unobserved infection times of a selection of VL cases, and indicates that the data does contain sufficient information to constrain the probable infection times of some cases, as the posterior distributions differ significantly from the prior distributions (i.e. from the prior for the incubation period distribution) for some cases.

Snapshots of one inferred transmission tree in the part of the south-east cluster of paras shown in Fig. 5 in the main text at different time points are shown in Figure S14. These show that while asymptomatic infection (and therefore subsequent immunity) was initially mostly clustered around VL cases, by December 2005 it was widespread and by December 2009 had saturated in this part of the study population. The relatively high uncertainty in infection sources can be seen in the different pattern of infectors and longer inferred transmission distances in this single tree (from one sample from the joint posterior distribution of model parameters and missing data) than the consensus tree in Fig. 5 in the main text.

**Fig. S12.**
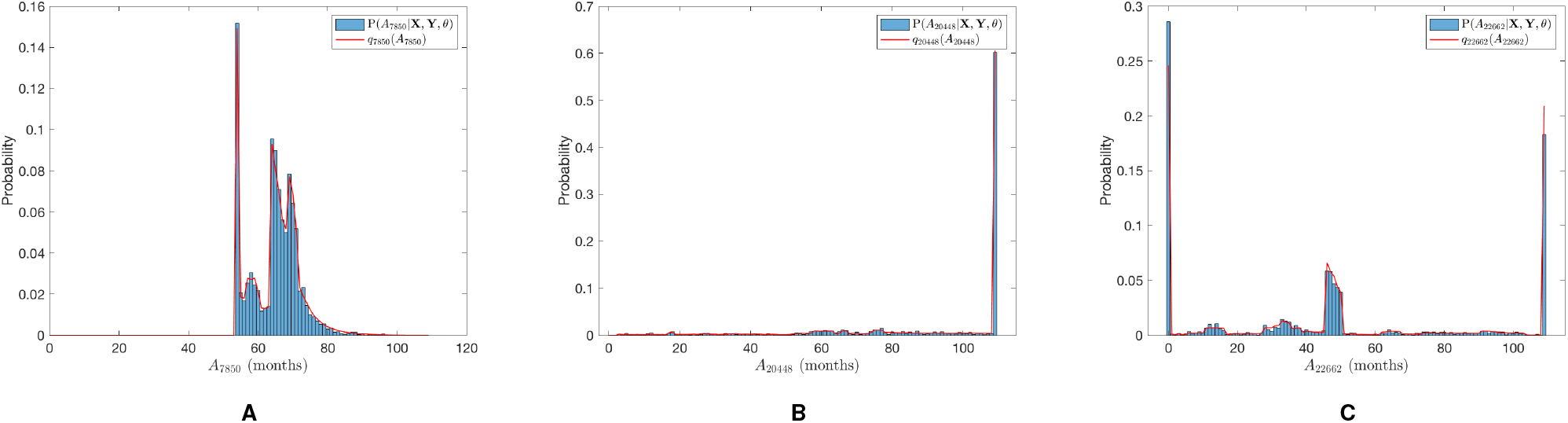
Posterior distributions of the unobserved asymptomatic infection times of a selection of individuals (blue) and their final asymptomatic infection time proposal distributions (red). Note that asymptomatic “infection” in months 0 and *T* + 1 = 109, represent asymptomatic infection before the study and no asymptomatic infection before the end of the study, respectively. (A) Individual who migrated into a house with an active VL case from outside the study area in month 53 and therefore had a high initial probability of asymptomatic infection, followed by further peaks in asymptomatic infection risk in months 64 and 69 with the PKDL and VL onsets of two other household members in months 63 and 68 respectively. (B) Individual born in month 2 with a high probability of having avoided asymptomatic infection for the duration of the study. (C) Individual who was 23-years-old at the start of the study with a moderately high risk of having been asymptomatically infected before the study and a small peak in asymptomatic infection risk when a fellow household member had VL onset in month 45.

**Fig. S13.**
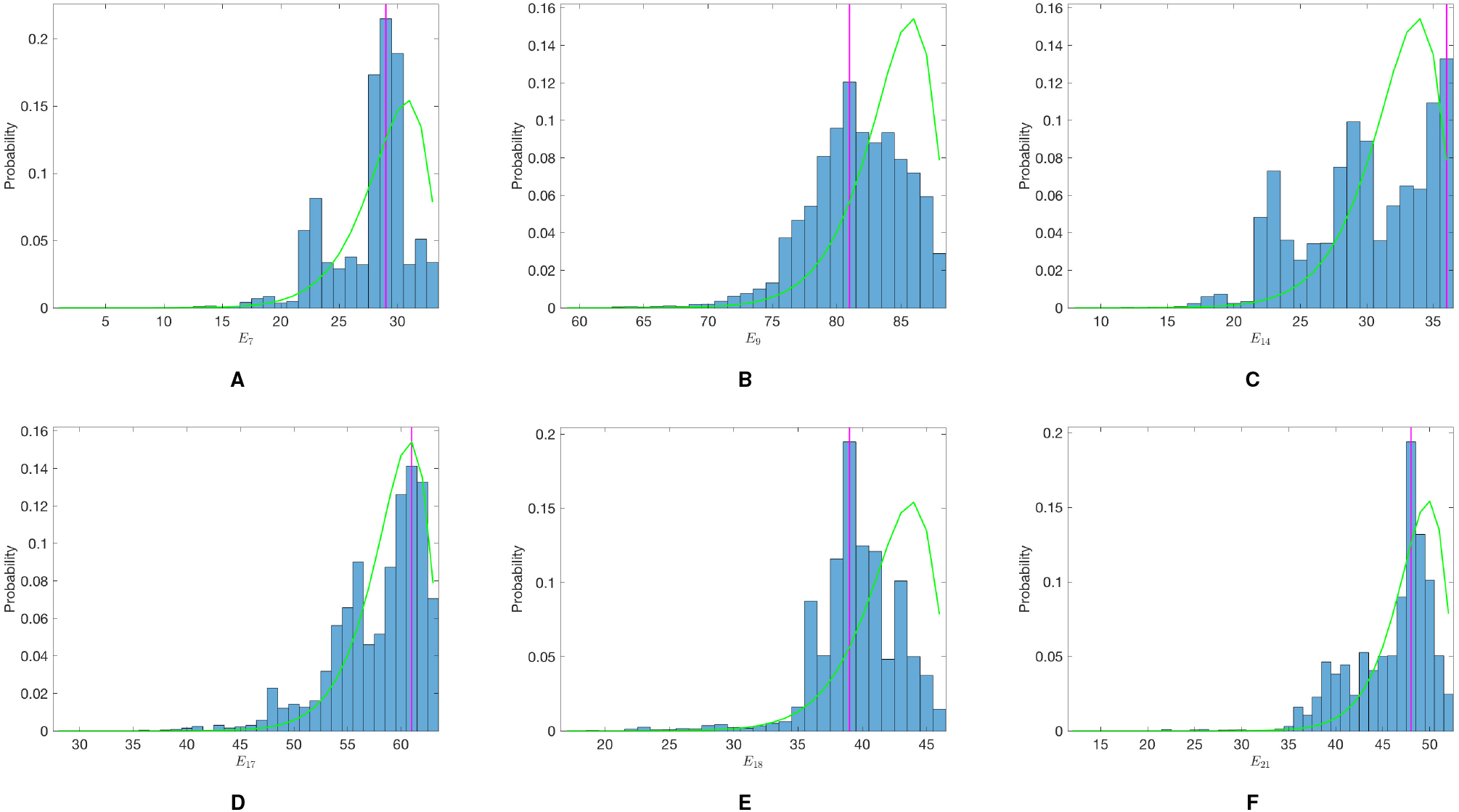
Posterior distributions of the unobserved infection times of a selection of VL cases with known onset times (blue histograms), with prior distributions (green lines) and modes (magenta lines).

**Fig. S14.**
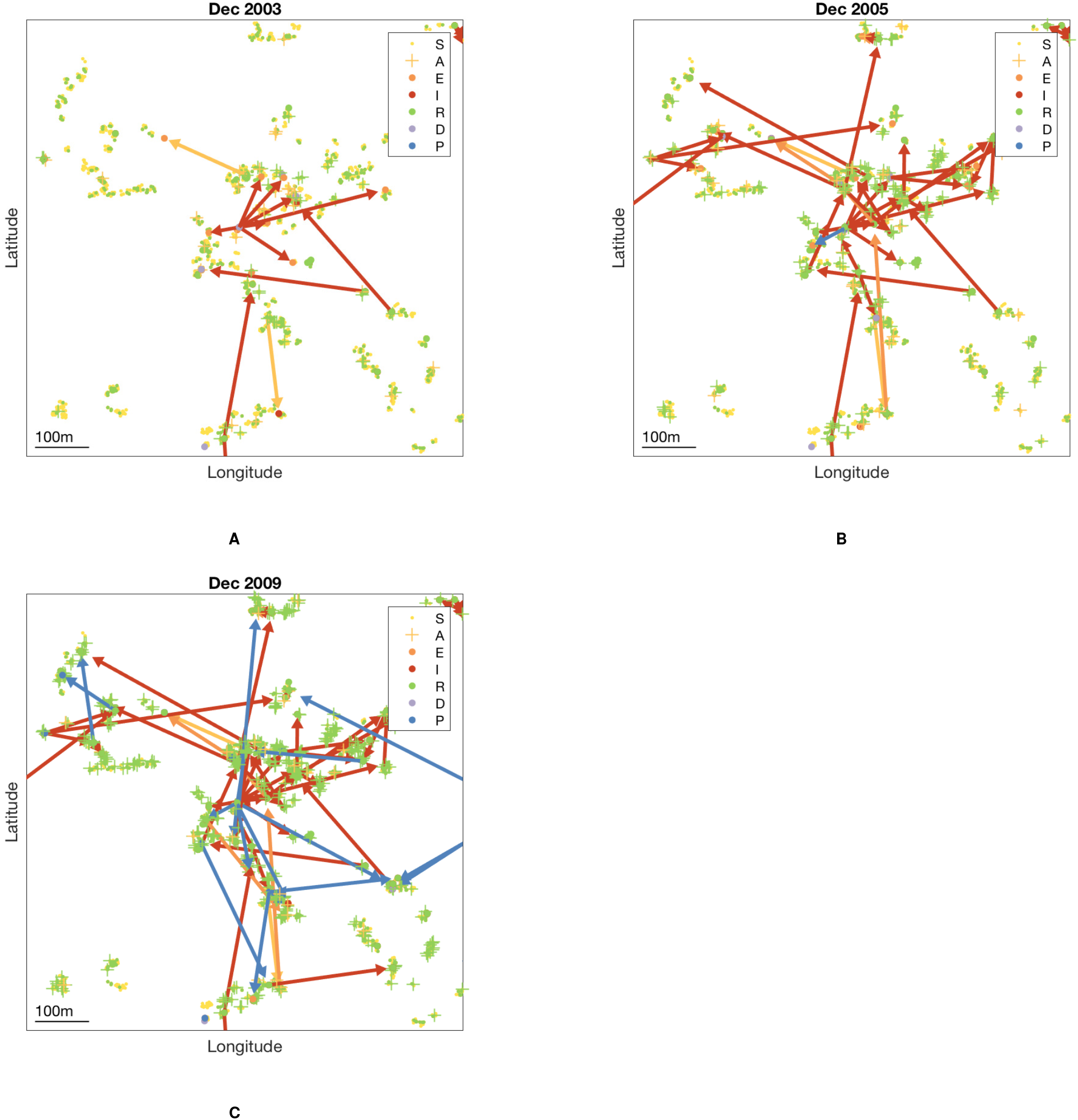
Inferred transmission tree in part of the south-east cluster of villages at different stages of the epidemic — (A) Dec 2003, (B) Dec 2005, and (C) Dec 2009 — from a single sample from the joint posterior distribution of model parameters and missing data. Model parameters chosen close to the posterior mode (*β* = 1.9 mnth^*−*1^, *α* = 114 m, = 5.6 *×* 10^*−*4^ mnth^*−*1^, *δ* = 0.100 mnth^*−*1^, *p* = 0.33). Dots show susceptible individuals/symptomatic cases and crosses asymptomatic individuals, and are coloured by infection state (see key). Arrows show the source of infection for each VL case infected up to that point in time and are coloured by type of infection source. GPS locations of individuals are jittered slightly so that individuals from the same household are more visible. An animated version showing all months is provided in SI movie S2.

**Table S5.** Para-level populations and VL and PKDL incidence 2002–2010

| Para | Cluster | Total no. individuals | Mean population | Total VL cases | Total PKDL cases | Mean VL incidence (cases/10,000/yr) | Mean PKDL incidence (cases/10,000/yr) |
| --- | --- | --- | --- | --- | --- | --- | --- |
| 1 | NW | 1398 | 1219 | 120 | 11 | 109 | 10 |
| 2 | NW | 1650 | 1376 | 46 | 3 | 37 | 2 |
| 3 | NW | 998 | 830 | 18 | 3 | 24 | 4 |
| 4 | NW | 2185 | 1768 | 197 | 35 | 124 | 22 |
| 5 | NW | 934 | 774 | 15 | 2 | 22 | 3 |
| 6 | NW | 1640 | 1363 | 94 | 22 | 77 | 18 |
| 7 | SE | 604 | 493 | 42 | 7 | 95 | 16 |
| 8 | SE | 585 | 496 | 8 | 0 | 18 | 0 |
| 9 | SE | 969 | 809 | 33 | 6 | 45 | 8 |
| 10 | SE | 701 | 595 | 17 | 3 | 32 | 6 |
| 11 | SE | 1300 | 1080 | 28 | 9 | 29 | 9 |
| 12 | SE | 2807 | 2388 | 102 | 25 | 47 | 12 |
| 13 | SE | 933 | 816 | 36 | 7 | 49 | 10 |
| 14 | SE | 446 | 391 | 23 | 3 | 65 | 9 |
| 15 | SE | 660 | 574 | 15 | 3 | 29 | 6 |
| 16 | NW | 905 | 762 | 26 | 9 | 38 | 13 |
| 17 | NW | 2080 | 1764 | 75 | 13 | 47 | 8 |
| 18 | NW | 3212 | 2653 | 61 | 11 | 26 | 5 |
| 19 | NW | 774 | 647 | 62 | 18 | 107 | 31 |
| Total |  | 24781 | 20798 | 1018 | 190 | 54 | 10 |

NW = north-west, SE = south-east.

**Fig. S15.**
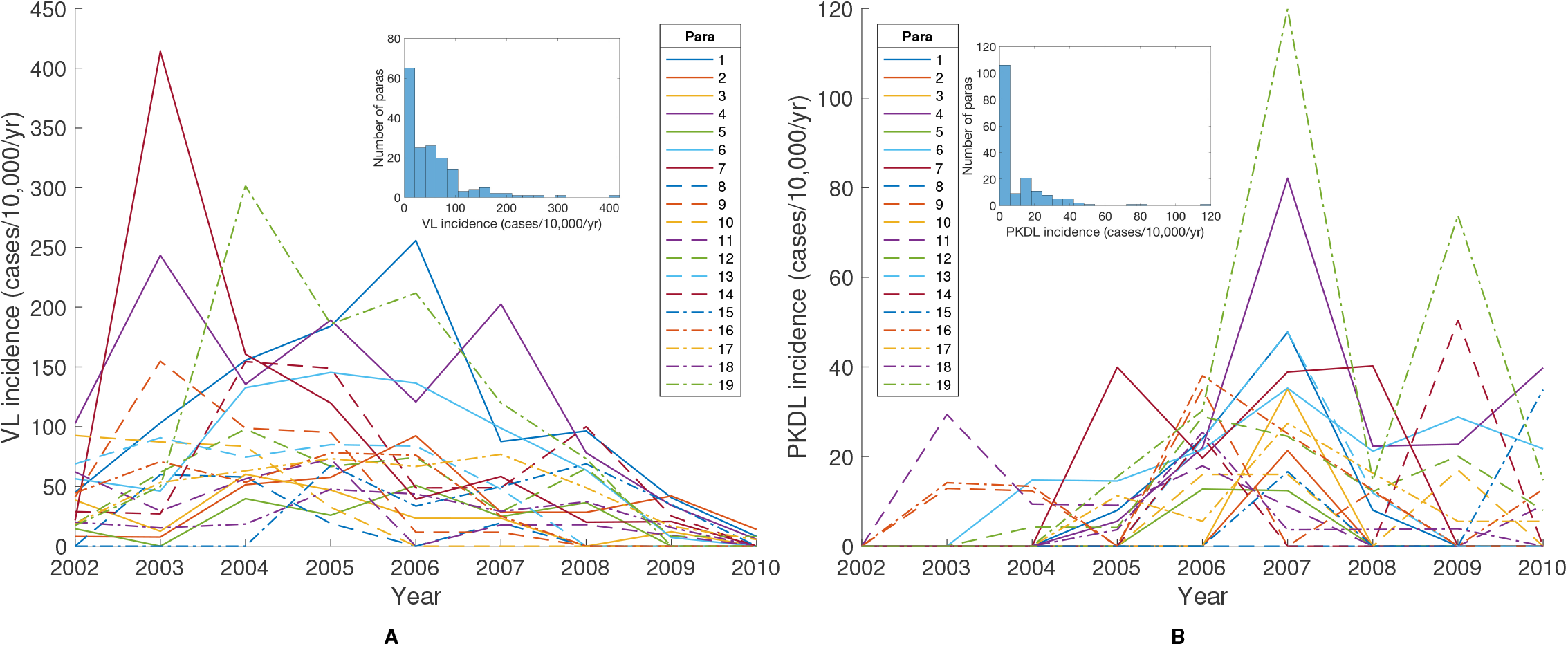
Para-level annual incidence of (A) VL and (B) PKDL. Insets: distributions of annual incidences.

**Fig. S16.**
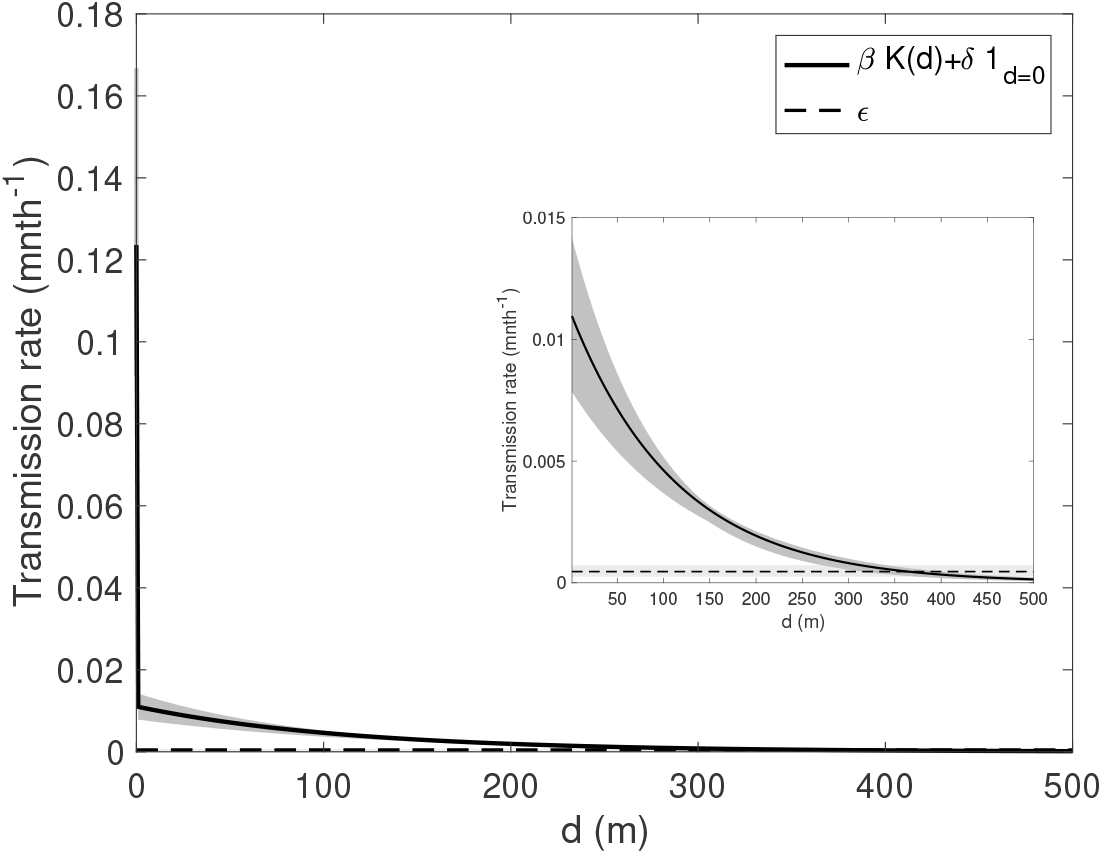
Estimated spatial transmission kernel showing decrease in infection risk with distance from an infectious individual. Transmission risk per month, *βK*(*d*), shown as a function of distance *d* away from a VL case (solid black line = mode, dark grey shaded region = 95% CI). Background transmission rate,, for unexplained infections also shown for comparison (dashed black line = mode, light grey shaded region = 95% CI). Inset: enlarged view of the part of the transmission kernel outside the household of the VL case.

**Fig. S17.**
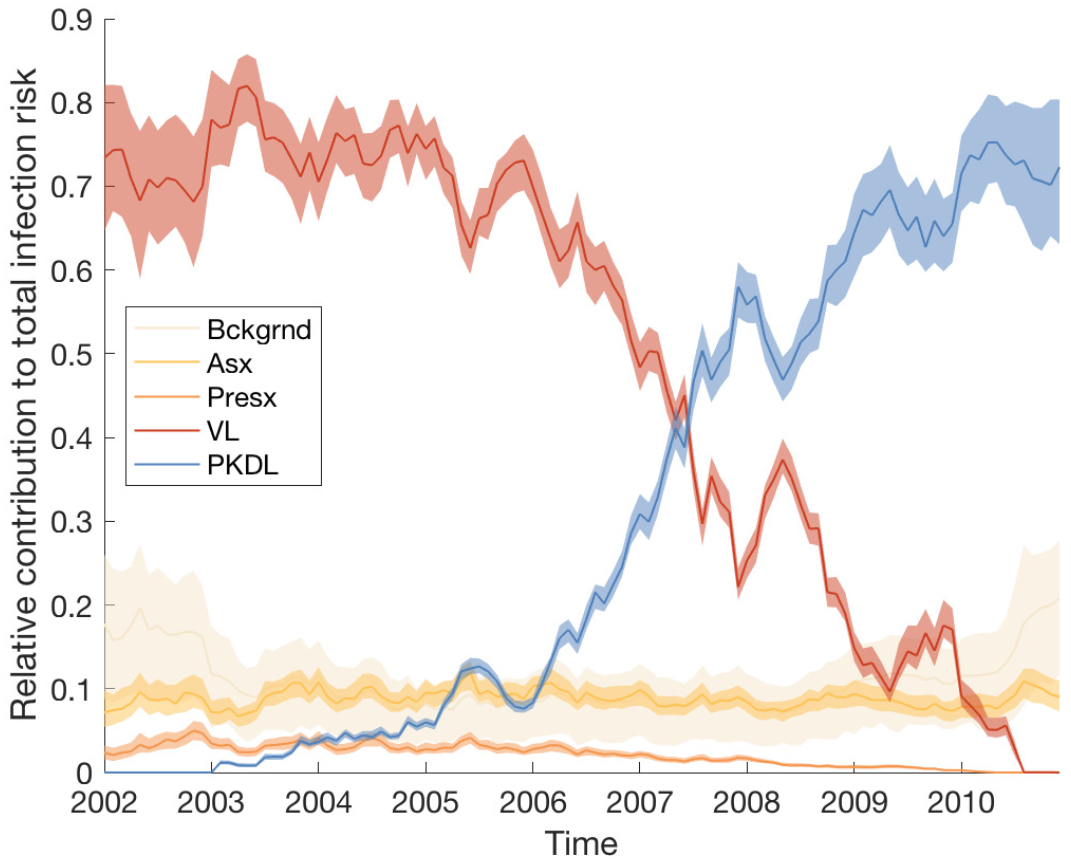
Relative contributions of background transmission, asymptomatic individuals, pre-symptomatic individuals, VL cases and PKDL cases to the total risk of new infections.

**Fig. S18.**
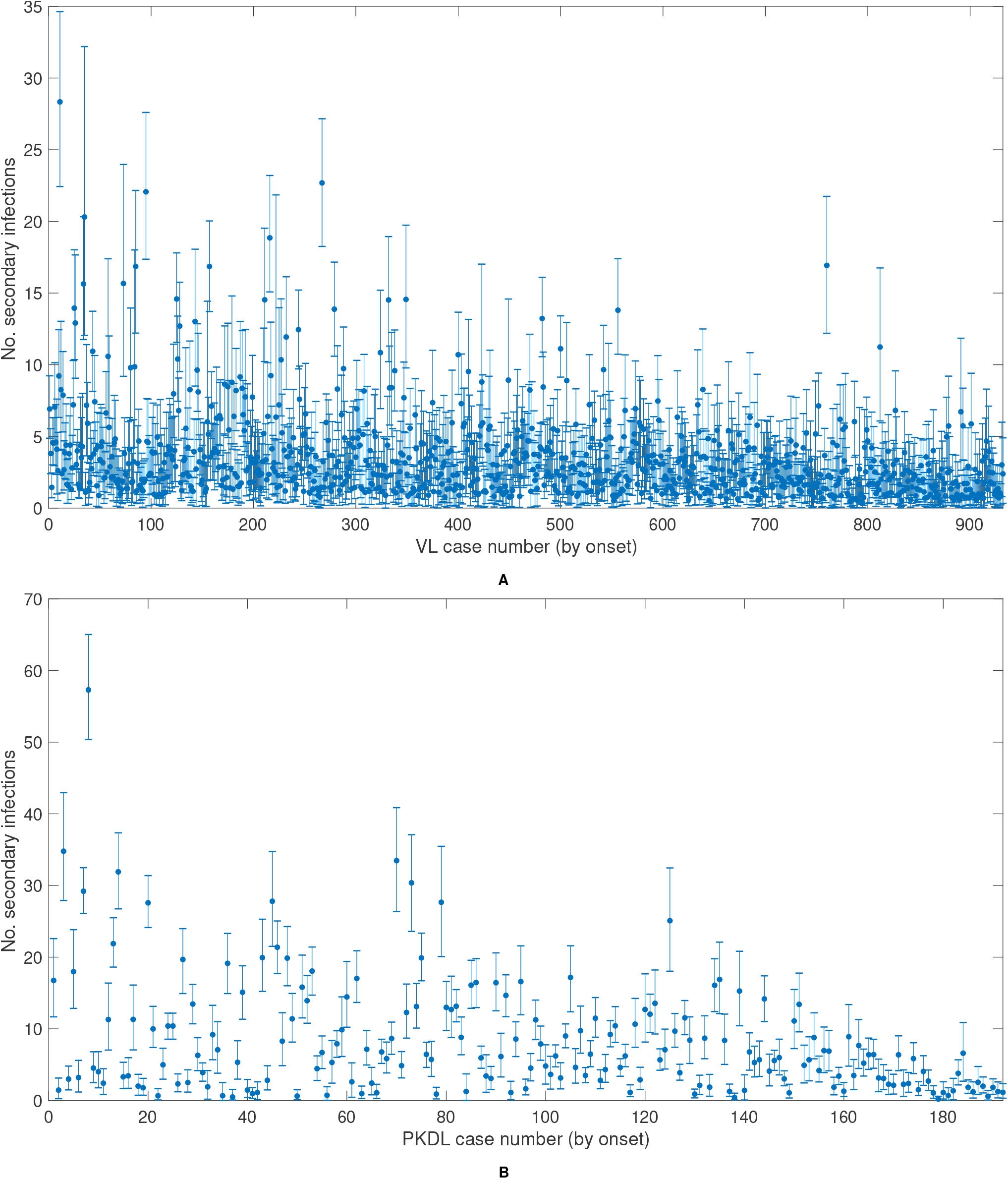
Number of secondary infections generated by (A) each VL case and (B) each PKDL case. Dots show mean reproduction number, bars show 95% CI (2.5–97.5% quantile).

**Fig. S19.**
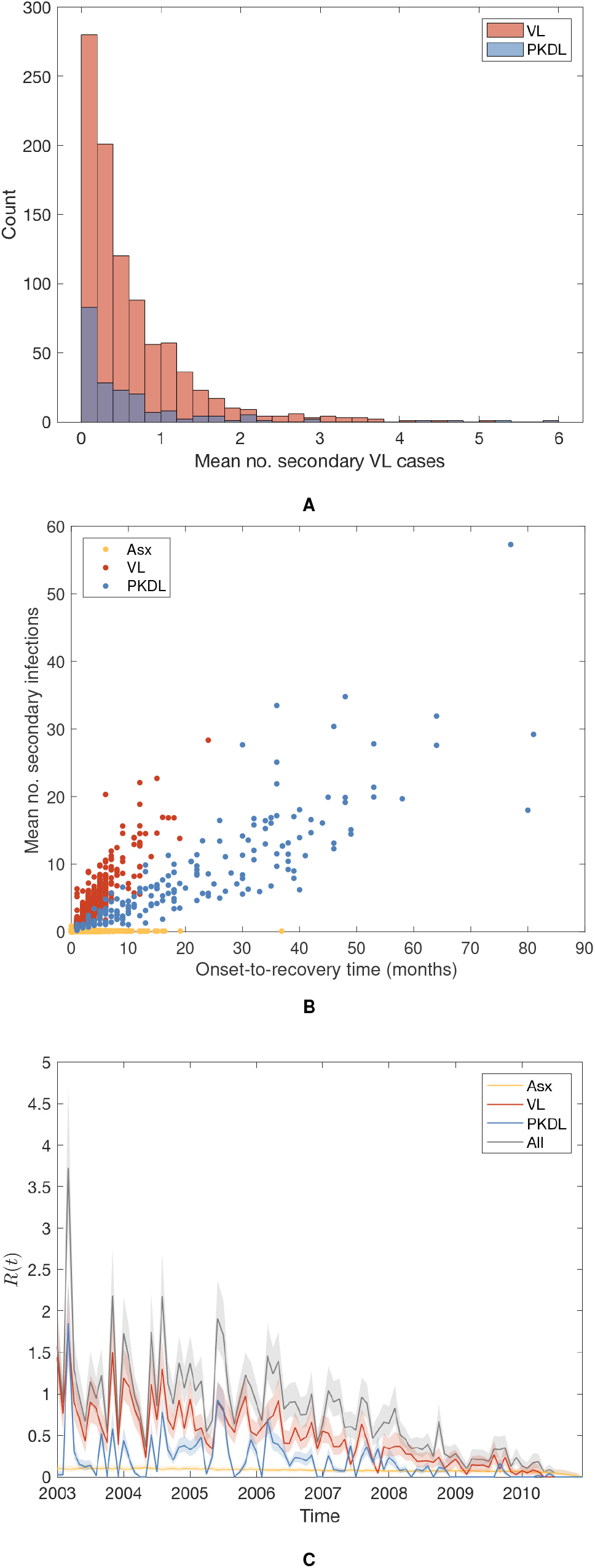
(A) Distributions of mean numbers of secondary VL cases per VL case and PKDL case. (B) Relationship between mean number of secondary infections and onset-to-recovery time for VL and PKDL cases and infection-to-recovery time for asymptomatic individuals. (C) Effective reproduction number *R*(*t*) with contributions from asymptomatic individuals, VL and PKDL cases. Solid lines show medians, and shaded bands 95% CIs.

**Fig. S20.**
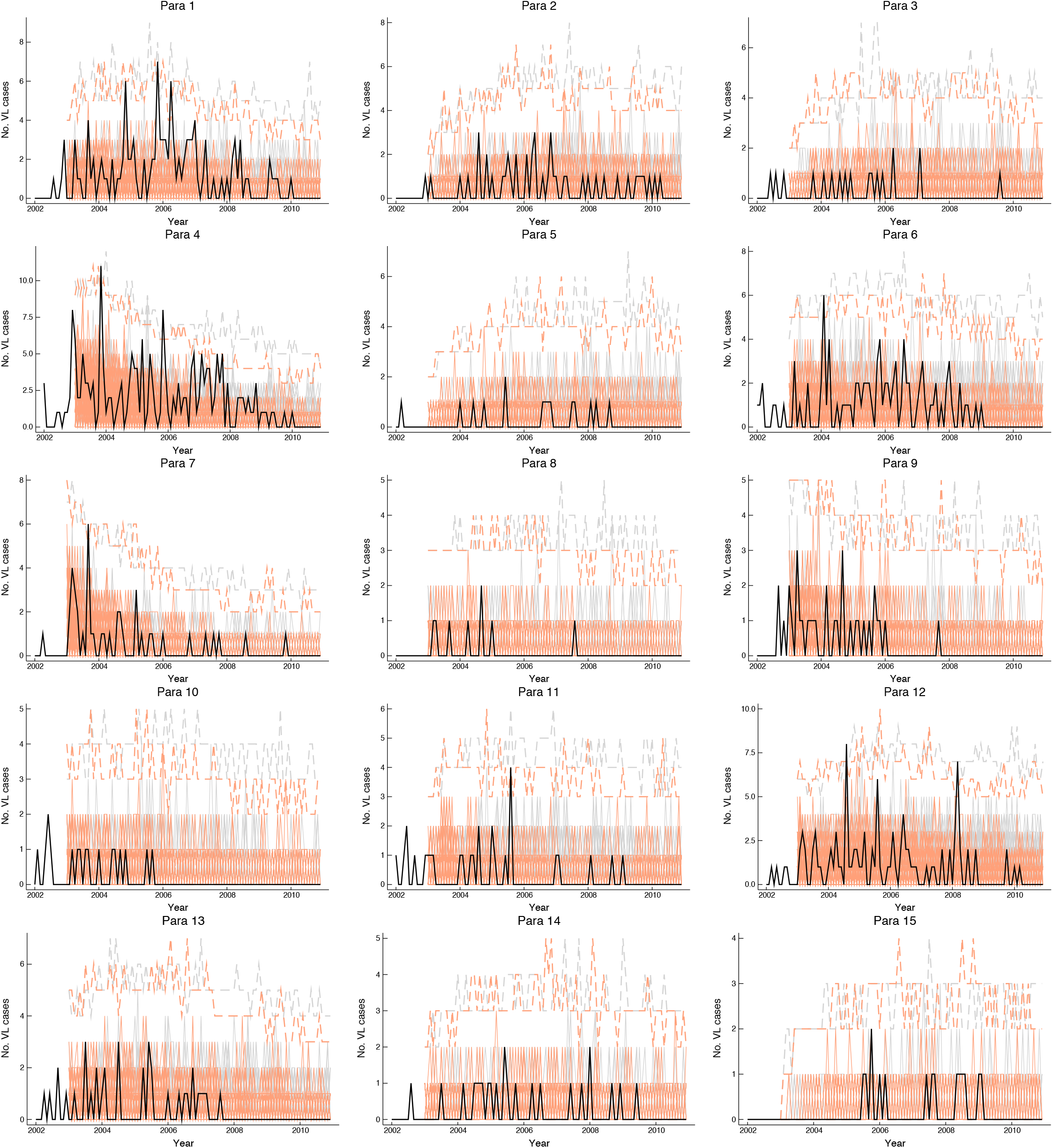
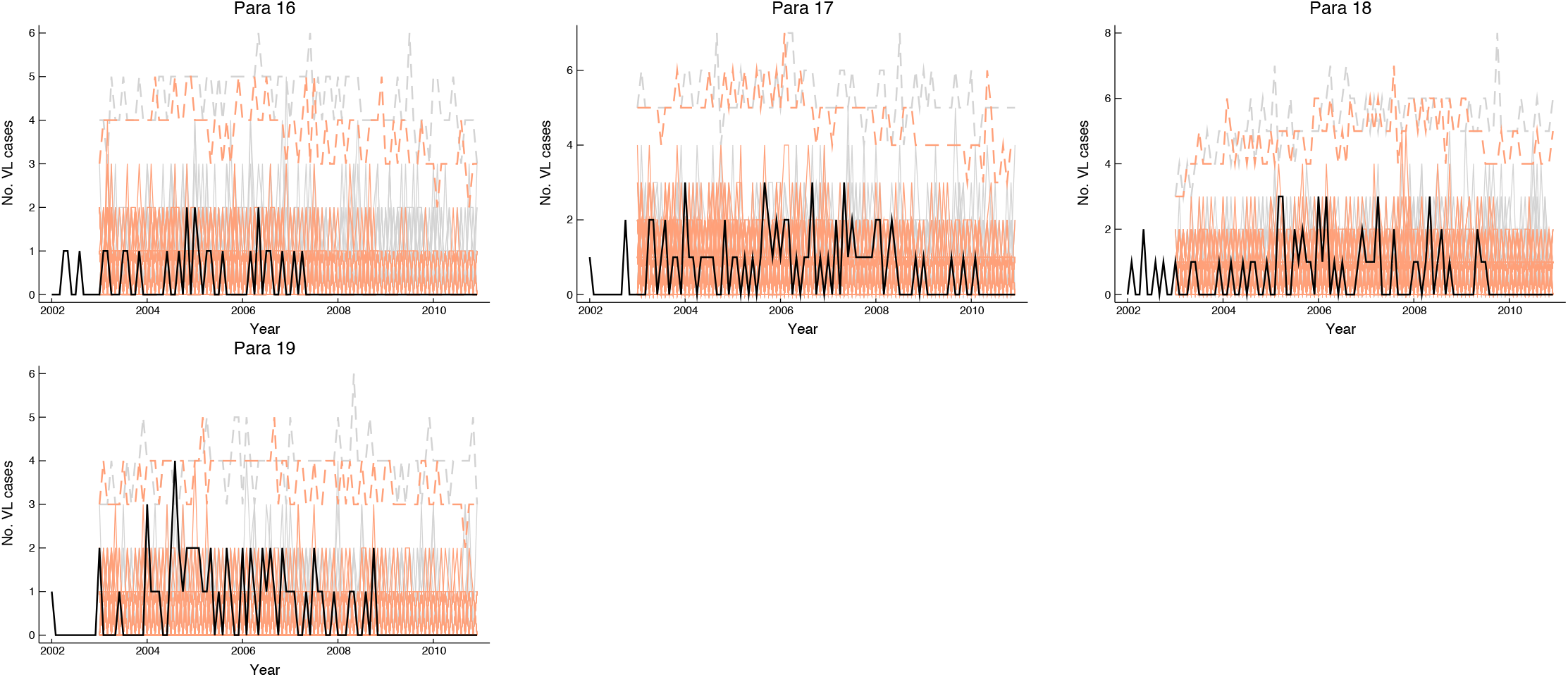
Para-level epidemic curves simulated with the individual-level spatiotemporal transmission model starting from January 2003 (by which point all but one para had had at least 1 VL case since January 2002) under normal conditions (solid grey lines) (17% of VL cases develop PKDL and 0.3% of asymptomatic individuals develop PKDL) and with complete prevention of PKDL (solid pink lines). Solid coloured lines show 100 randomly chosen simulations from 10,000 simulations (100 samples of the model parameters and initial infection statuses from the posterior distribution each simulated 100 times), solid black lines show observed incidence of VL, dashed lines show the minimums and maximums of all the simulations at each time point.

**Table S6.**
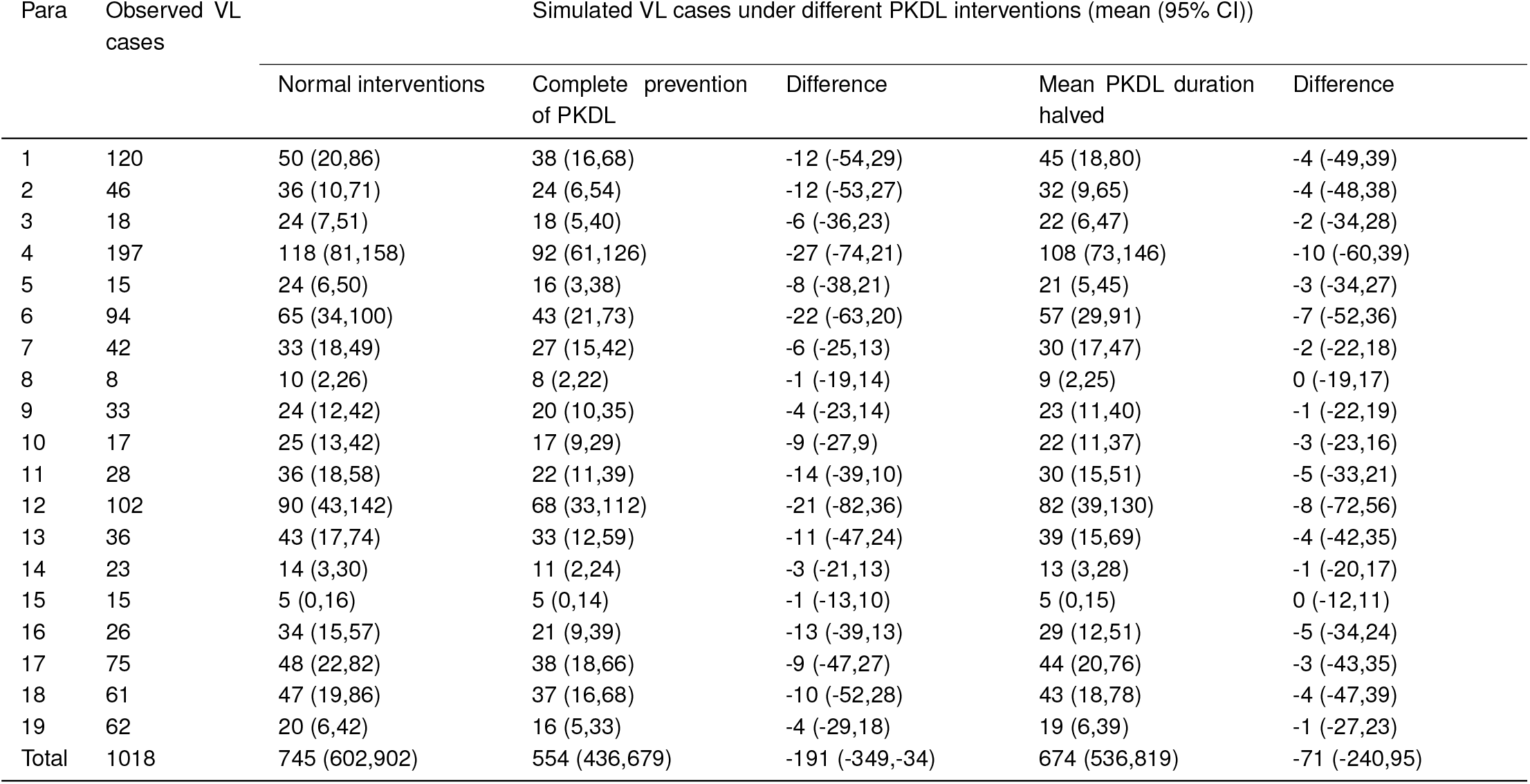
Observed and simulated numbers of VL cases in each para for different PKDL interventions 2002–2010

**Movie S1. Animation of the evolution of the most likely transmission tree in Fig. 3 in the main text over the course of the epidemic from January 2003 (***t* = 13**) to December 2010 (***t* = 108**). Dots show individuals coloured by their infection state (see key). Arrows show the most likely source of infection for each case over 1000 sampled transmission trees, coloured by type of infection source and shaded according to the proportion of trees in which that individual was the most likely infector (darker shading indicating a higher proportion)**.

**Movie S2. Animation of the transmission tree shown in Fig. S14 from a single sample from the joint posterior distribution of model parameters and missing data. Dots, crosses and arrows as in Fig. S14**.

